# Optimal Stimulation Sites and Networks for Deep Brain Stimulation of the Fornix in Alzheimer’s Disease

**DOI:** 10.1101/2022.09.08.22279028

**Authors:** Ana Sofía Ríos, Simón Oxenford, Clemens Neudorfer, Konstantin Butenko, Ningfei Li, Nanditha Rajamani, Alexandre Boutet, Gavin J.B. Elias, Jurgen Germann, Aaron Loh, Wissam Deeb, Fuyixue Wang, Kawin Setsompop, Bryan Salvato, Leonardo Almeida, Kelly D. Foote, Robert Amaral, Paul B. Rosenberg, David F. Tang-Wai, David A. Wolk, Anna D. Burke, Stephen Salloway, Marwan N. Sabbagh, M. Mallar Chakravarty, Gwenn S. Smith, Constantine G. Lyketsos, Michael S. Okun, William S. Anderson, Zoltan Mari, Francisco A. Ponce, Andres M. Lozano, Andreas Horn

**Author notes:** **Corresponding Author** Andreas Horn, MD, PhD, Associate Professor of Neurology, Center for Brain Circuit Therapeutics, Department of Neurology, Brigham and Women’s Hospital, MA, USA.

## Abstract

Deep brain stimulation (DBS) to the fornix is an investigational treatment option for patients with mild Alzheimer’s Disease. Outcomes from randomized clinical trials have shown that cognitive function improved in some patients but deteriorated in others. One reason could be variance in electrode placement leading to differential engagement of neural circuits. To investigate this, we analyzed a multi-center cohort of 46 patients with DBS to the fornix. Using normative structural and functional connectivity data, we demonstrate that stimulation of the circuit of Papez and stria terminalis robustly associated with cognitive improvement (R = 0.45, p = 0.031). On a local level, the optimal stimulation site resided at the direct interface between these structures (R = 0.33, p = 0.016). Finally, modulating specific distributed brain networks related to memory accounted for optimal outcomes (R = 0.38, p = 0.006). Findings were robust to multiple cross-validation designs and may now define an optimal network target which could subsequently guide refinement of DBS surgery and programming.

## Introduction

Alzheimer’s Disease (AD) is the most common neurodegenerative disease and the fifth leading cause of death in adults older than 65 years with an increasing total healthcare burden currently above $300 billion per year in the US^1^, thus, finding effective treatment options for AD has great socioeconomic relevance. The pathophysiology of AD is associated with amyloid beta (Aβ) protein depositions, phosphorylated tau protein tangles, neuronal and synaptic loss, as well as neurotransmission deficits. Neuronal loss, in particular, results in gross cerebral atrophy with a predilection for structures implicated in memory function, including the Papez circuit^2^ and components of the default mode network (DNM)^3,4^. Clinically, these neurodegenerative processes manifest as disturbances in memory, language and executive functions, as well as progressive loss of day to day functional abilities^5^. Given the well described patterns of Aβ^6^ and tau depositions^7^, these have been the target of therapeutic efforts for over 20 years. However, these efforts have been without significant success leading to a range of alternative approaches to modify the disease. One promising approach has been based on observed network alterations throughout the brain^3,4,8^, such as decreased connectivity in precuneus, parahippocampal gyrus, thalamus and post central gyrus^9^, as well as white matter disruption^10^, in addition to Aβ and tau patterns^5^, thus conceptualizing AD as a “circuitopathy”^11^.

Deep brain stimulation (DBS) has been shown to successfully alleviate symptoms in circuit disorders of the human brain such as Parkinson’s Disease^12^, Essential Tremor^13^, and – more recently – obsessive-compulsive disorder^14^ and other neuropsychiatric disorders^15^. In addition to evidence of sensory stimulus producing gamma entrainment and subsequent reduction of amyloid pathology and improvement in spatial and recognition memory in an AD-mice model^16,17^, research has grown that investigates neuromodulation in the treatment of AD. DBS to the fornix (fx-DBS) has emerged as an investigational treatment targeting associated circuit disruptions with the aim of modulating associative and limbic networks that subserve memory function, and most specifically the Papez’ circuit^18^. In addition to evidence of fornix atrophy in mild cognitive impairment (MCI)^19^, as a diagnostic or prognostic marker in AD^20–22^, and as an essential component of memory formation and consolidation^21^, its potential utility as a DBS target was considered after a serendipitous observation of flashback-like episodes during DBS of the hypothalamic region in a patient with morbid obesity^23^. Although the occurrence of such memory events had been reported previously in the context of temporal lobe stimulation^24^, the observation of flashback phenomena after hypothalamic region stimulation, in proximity to limbic structures such as the fornix, led to a first series of six AD patients receiving fx-DBS^25^. While alternative DBS target regions including the ventral capsule/ventral striatum^26^ and the nucleus basalis of Meynert^27^ have been proposed, the fornix has become the most studied region with over 101 patients having undergone this intervention to date^28^. There are now completed phase I^25^ and II^29^ clinical trials (NCT00658125, NCT01608061), as well as an ongoing international randomized-controlled trial (Advance II, NCT03622905)^29^. In addition, recent studies investigated the neural substrates underlying memory flashbacks^30,31^ and autonomic response^32^ reported in this patient population.

Fx-DBS has been hypothesized to impact circuits by modulating glucose metabolism impairment in temporal and parietal regions, and there is evidence of hippocampal volume increase in mildly affected AD patients after 6 months of stimulation^25^. Nevertheless, the clinical benefits of fx-DBS remain unproven with promising outcomes for some patients, but no benefit for others. Age has emerged as a possible treatment effect modifier in the ADvance trial. Here, among individuals in the early-on arm during phase 1 (but not in phase 2), participants below the age of 65 worsened on the ADAS-cog13 significantly more than older participants^33^.

A competing explanation for differences in clinical outcomes across patients could be *variance in electrode placement*, as demonstrated across multiple disorders treated with DBS^12,14,34–36^. The effect of lead location could be stronger in investigational DBS targets where the exact target is not yet precisely defined (leading to more variance in placement) and the neural substrates driving clinical outcome remain poorly understood^14,34,36,37^.

Growing interest in the therapeutic potential of fx-DBS in AD has led to a sizable cohort of patients receiving electrode implantation with this target in recent years. In the present study we leveraged a unique, multi-centric, large dataset (N = 46) of patients treated with fx-DBS, to investigate variability in DBS electrode placement on three levels: i) effects of focal electric fields of stimulation on white matter tracts traversing the stimulation volumes ii) optimal stimulation sites on a localized voxel level, and iii) impact of fx-DBS on distributed whole-brain functional networks. We do so by applying a state-of-the-art DBS electrode localization method^35^ and a subsequent DBS fiber filtering^38^ and network mapping approaches^1^.

## Results

### Patient demographics and clinical results

We performed a post hoc analysis on a series of 46 patients (mean age: 67 ± 7.9 years, 22 females) with mild AD (ADAS-cog 11: 12-24 points; CDR of 0.5 or 1.0) who underwent bilateral DBS (electrode type: Medtronic 3387, Medtronic, Minneapolis, MN) targeting the fornix region across seven international centers between 2007 and 2019^25,29^, following a standardized stimulation protocol (see figure S1 for patient selection flow, tables S1 and S2 for inclusion/exclusion criteria, and table S3 for patients scores). All patients received DBS at a frequency of 130 Hz and pulse width of 90 ms. AD patients had an ADAS-cog 11 score of 18.5 ± 5.6 (mean ± SD) at baseline and 23.6 ± 10 one year after stimulation (−38.6 ± 48.8 % change). In each patient, electrode placement was reconstructed using the revised pipeline of Lead-DBS (www.lead-dbs.org^35^). Electrode localization confirmed accurate placement within the ventral diencephalon in all patients (Figure S2). However, differences in electrode placement could be observed across patients: 73/92 active contacts featured a radius ≤ 2 mm to the closest voxel of the fornix^39^. Similarly, 85/92 active contact centers were located ≤ 2 mm apart from the closest voxel of the Bed nucleus of Stria Terminalis (BNST)^39^.

To investigate differential DBS effects on structures more deliberately, electric fields were estimated for the chronic DBS stimulation parameters using a finite element modeling (FEM) approach as implemented in Lead-DBS^35^. Based on the electric field magnitude (E-field), DBS effects were investigated on the white matter (*DBS fiber filtering*^38^, Figure 1A), focal (*DBS Sweetspot Mapping*^40^, Figure 1B), and distributed network (*DBS network mapping*^12^, Figure 1C) level, results were then cross-validated using Leave-one-patient-out and k-fold (3, 5, 7 and 10-fold) designs. For fiber filtering and network mapping, normative connectivity data estimated in healthy subjects was used to define streamlines and regions of interest in this cohort (see table S4 for underlying data on normative connectomes).

**Figure 1:**
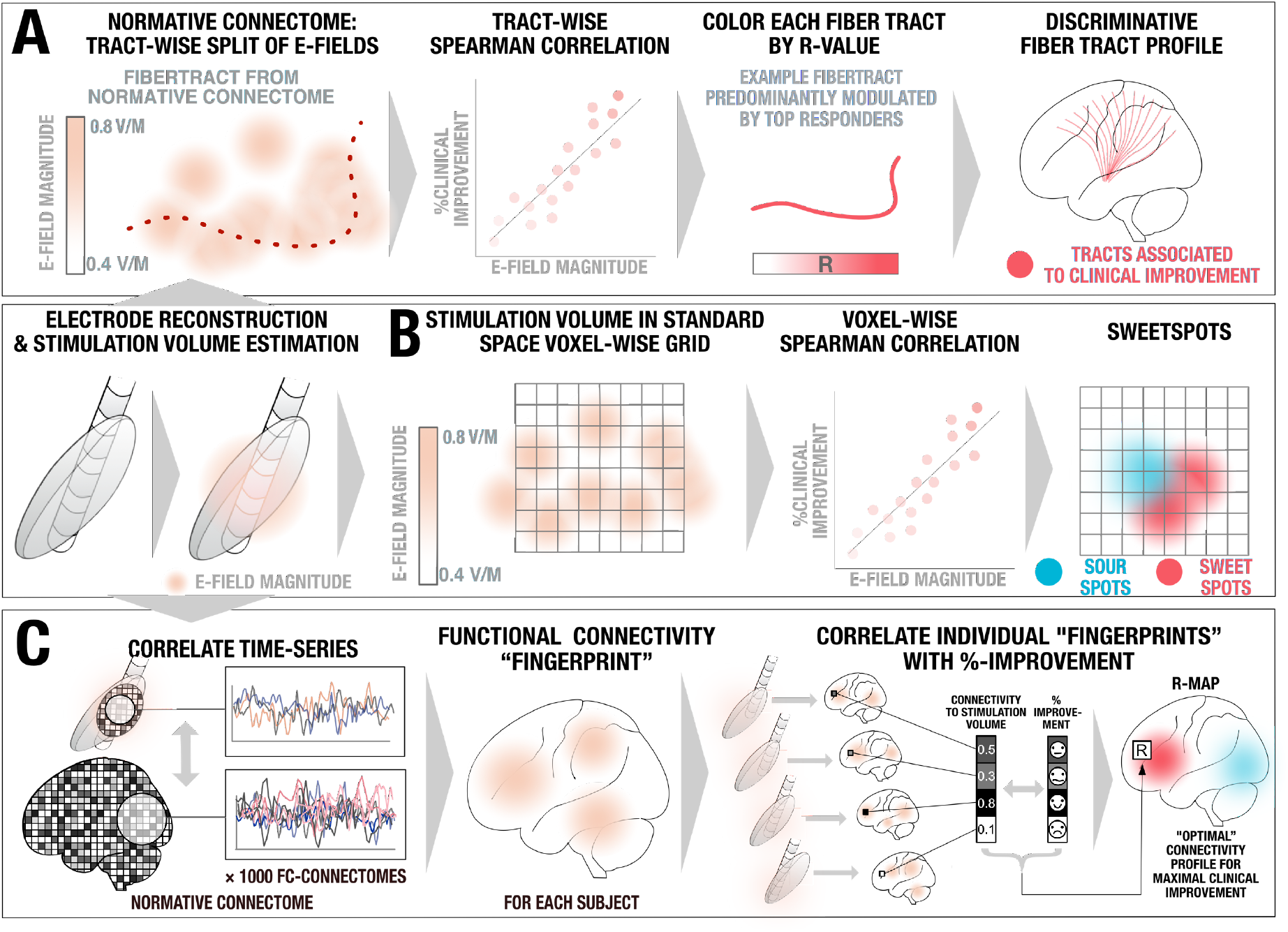
Overview of the three methods applied. A pre-requisite to run these analyses is to reconstruct the electrode trajectory and localization to then estimate the stimulation volume following the finite element method (FEM). A) DBS fiber filtering. Stimulation volumes as E-fields were pooled in standard space and overlaid on an ultra-high resolution normative connectome^41^. Peak E-field magnitudes along each tract were aggregated for each stimulation volume and Spearman rank-correlated assigning weights to each tract based on respective clinical outcomes. B) DBS Sweetspot mapping. For each voxel, the E-field vector magnitudes and clinical outcomes were Spearman rank-correlated, leading to a map with positive and negative associations (sweet and sour spots). C) DBS network mapping. Seeding BOLD-signal fluctuations from each E-field in a normative functional connectome consisting of rs-fMRI scans from 1,000 healthy participants^42^ yielded a functional connectivity “fingerprint” map for each patient. Maps were then Spearman rank-correlated with clinical improvement in a voxel-wise manner to create an R-map model of optimal network connectivity.

### Tracts associated with optimal DBS response (DBS Fiber Filtering)

As the core analysis of this study, we determined the stimulation of *which fiber tracts* were associated with maximal clinical improvement. This analysis should be seen as the main analysis of the present work since i) the fornix constitutes a network target aiming to modulate distributed network activity within the circuit of Papez, ii) the target is a white-matter structure readily identifiable by structural imaging and tractography, and iii) tractography could be used to define tract-targets in prospective clinical trials, as has been done previously^43^. We applied the DBS fiber filtering approach, introduced in^38^ and methodologically generalized for use with E-fields in^44^. While the method has led to robust results that were predictive across DBS cohorts and surgeons in multiple reports and indications^14,34,44^, it should still be considered a novel approach and fiber filtering results hence warrant multiple levels of validations. To do so, patients were first randomly split into a training (N = 28) and test (N = 18) cohort. We then performed DBS fiber filtering on the training set using an ultra-high resolution normative connectome calculated from a 760 µm resolution whole-brain diffusion scan^41^ to identify a set of white matter streamlines connected to the stimulation volume (thresholded E-field following Astrom et al^45^) of each patient and correlated to optimal clinical outcomes informed by ADAS-cog 11 scores (Figure 2A, see table S5 for fiber filtering parameters).

**Figure 2.**
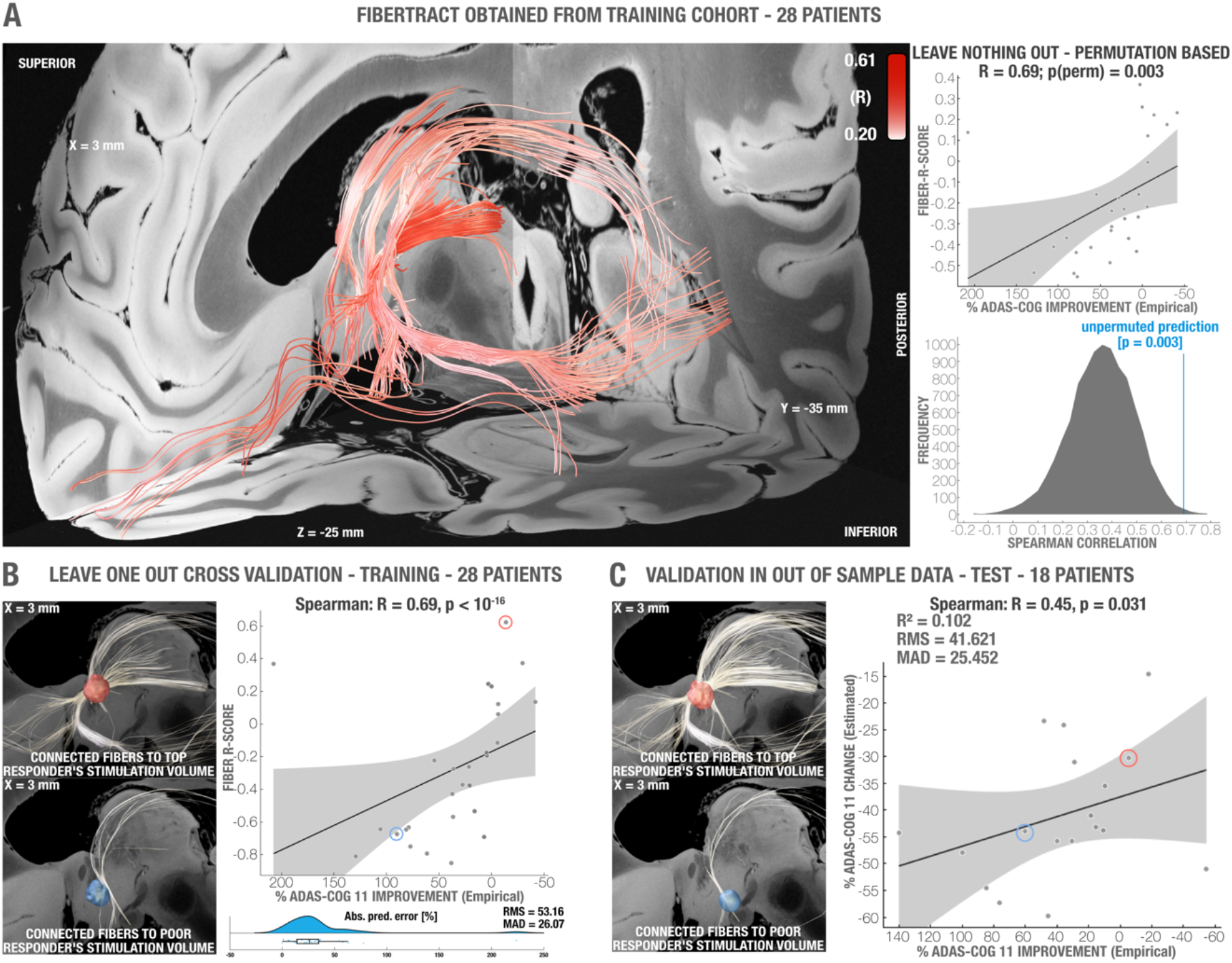
Validation of tract models predictive of clinical improvements as evaluated using ADAS-Cog 11. A) Left: Optimal set of tracts to be modulated as calculated from the entire training cohort (N = 28 subjects). Right: permutation analysis calculated on the entire training cohort. B) Top left: stimulation volume of a patient with top clinical improvement overlapping the tracts associated with optimal clinical improvements (calculated leaving out the subject, N = 28-1 = 27 subjects). Fibers displayed in white correspond to the portion of optimal fibers intersecting with the patient’s stimulation volume. Bottom left: Same analysis carried out with a poor-responding example patient. Right: Cross-validation within the training cohort using a leave-one-out design (top, R = 0.69 at p < 10^−16^) and within-fold analysis (bottom). The two example patients are marked in the correlation plot with circles. C) Optimal tracts calculated from the entire training cohort (as shown in panel A, N = 28) were used to cross-predict outcomes in N = 18 left out patients of the test cohort (R = 0.45, p = 0.031). Left: two example cases from the test cohort are shown, a top responding patient’s stimulation volume with corresponding connected (white) optimal fibers defined by the training cohort; and a poor-responding patient’s stimulation volume with corresponding connected (white) fibers. The two example patients are marked in the correlation plot with circles. Fiber tracts and example stimulation volumes were superimposed on slices of a 100-µm, 7T brain scan in MNI 152space^48^. Gray shaded areas represent 95% confidence intervals.

The tracts that accounted for optimal improvement in the training cohort followed the trajectory of the fornix, a parallel bundle ascending from the BNST which likely corresponds to the stria terminalis, as well as projections connecting to the anterior and ventral anterior nuclei of the thalamus, and an additional anterior, orbito-frontal projection. To validate this set of connections, we first cross-validated the model within the training cohort. To this end, the model was iteratively re-calculated in a leave-one-out design, each time estimating an individual patient’s improvement based on the streamlines defined in the remaining cohort (Spearman’s R = 0.69 at p < 10^−16^; Fig. 2B). The discrepancy with the actual improvement was quantified using the root means square (RMS = 53.16) and the median absolute deviation (MAD = 26.07, which is not as susceptible to outliers), shown on Figure S3. To further test robustness, we cross-validated the model within the training cohort using k-fold designs that again led to positive and significant correlations between predicted and empirical scores (3-fold: R = 0.52 at p = 0.002; 5-fold: R = 0.58 at p < 10^−16^; 7-fold: R = 0.65 at p < 10^−16^; 10-fold: R = 0.56 at p < 10^−16^, see Figure S3 for additional metrics); additionally, 1,000 permutations were computed for the training cohort, obtaining an R = 0.69 at p = 0.003 for non-permuted fibers (Figure 2A). This demonstrated high robustness of findings *within* the training cohort. Figure 2B shows two example patients of the *training cohort* with optimal (and correspondingly high E-field overlap with tract model calculated based on all but that one patient) and poor outcome (with minimal overlap), respectively.

Next, we used the fiber model calculated on the entire training cohort (N = 28) to estimate clinical outcomes in patients from the test cohort (N = 18), which had been left as a completely naïve hold-out set. This cross-cohort-prediction revealed a significant relationship (R = 0.45 at p = 0.031, R^2^ = 0.102, RMS = 41.621, MAD = 25.452; Figure 2C) indicating robustness of the generated model. It should be emphasized that for out-of-sample testing, we calculated the coefficient of determination R^2^ based on the sum of squared errors, and not by squaring the correlation coefficient^46^. Figure 2C again features two example patients – this time from the *test cohort* – with either optimal clinical outcome (and correspondingly maximal E-field overlap with the tract model calculated on the complete training cohort) or poor outcome (with minimal overlap), respectively.

As further evaluation, we calculated the predictive tract model based on the training-, test- and combined cohorts, separately. This allowed a direct comparison of results calculated in each cohort by visual inspection, and overlayed the identified bundle with structures of interest from atlases available in MNI space^39,47^ (Figure S4). Importantly, we ruled out that this set of connections does not simply represent the average connectivity site from electrodes but indeed a specific subset of connections *associated with clinical improvements*. This was confirmed by repeating the analysis after permuting improvement values across patients, which isolated different connections in each run (Figure S5), demonstrating the identified and robust set of connections specifically account for improvements following DBS.

As a final validation step, we carried out a leave-one-out cross validation across the *entire cohort* which led to an R = 0.66 at p < 10^−16^, RMS = 50.32, MAD = 33.23 between estimated fiber scores and empirical improvements. Further cross-validation k-fold designs led to similar results (3-fold: R = 0.44 at p = 0.002; 5-fold: 0.50 at p < 10^16^; 7-fold: R = 0.48 at p = 0.001; and 10-fold: R = 0.52 at p < 10^16^, Figure 5, see Figure S7 for additional metrics).

These analyses show robustness and predictive utility of tracts associated with optimal clinical outcomes across cohorts and may constitute a finding of great importance that could influence clinical practice (see discussion), especially with respect to guiding DBS programming after surgery. However, a practical clinical question before surgery is which target *coordinate* to use during surgical planning. To analyze this question, we carried out a voxel-wise mapping analysis to identify an optimal target sweet spot.

### Optimal stimulation site mapping (Sweetspot Analysis)

Sweetspot analysis revealed a consistently symmetric map across the two hemispheres with optimal stimulation sites located at the level of the anterior commissure (AC) extending into the descending columns of the fornix bilaterally (Figure 3). Non-linear flipping of stimulation volumes along the intercommissural plane (which doubles the N of correlations and would be sensible under the assumption of a symmetric DBS effect) led to a similar finding. Peak coordinates and centers of gravity of each cluster are given in table S6 for both analyses (see also Figure S6 for cluster center). The optimal stimulation site was located on the lateral and posterior portions of the columns of the fornix with peak R-values of −0.80 (sourspot) and 0.93 (sweetspot) with unflipped data and −0.66 (sourspot) and 0.77 (sweetspot) with flipped data. Note that these correlation coefficients should not be considered significant due to the mass-univariate (voxel-wise) design. Instead, spatial maps consisting of sweet- and sour-spots were cross-validated across the entire cohort in a leave-one-patient-out design, which led to significant results (R = 0.33 at p = 0.016, RMS = 50.60, MAD = 27.94). Further cross-validation designs led to similar results (3-fold: R = 0.27 at p = 0.037; 5-fold: R = 0.30 at p = 0.016; 7-fold: R = 0.39 at p = 0.005; 10-fold: R = 0.33 at p = 0.011, Figures 5 and S7).

**Figure 3.**
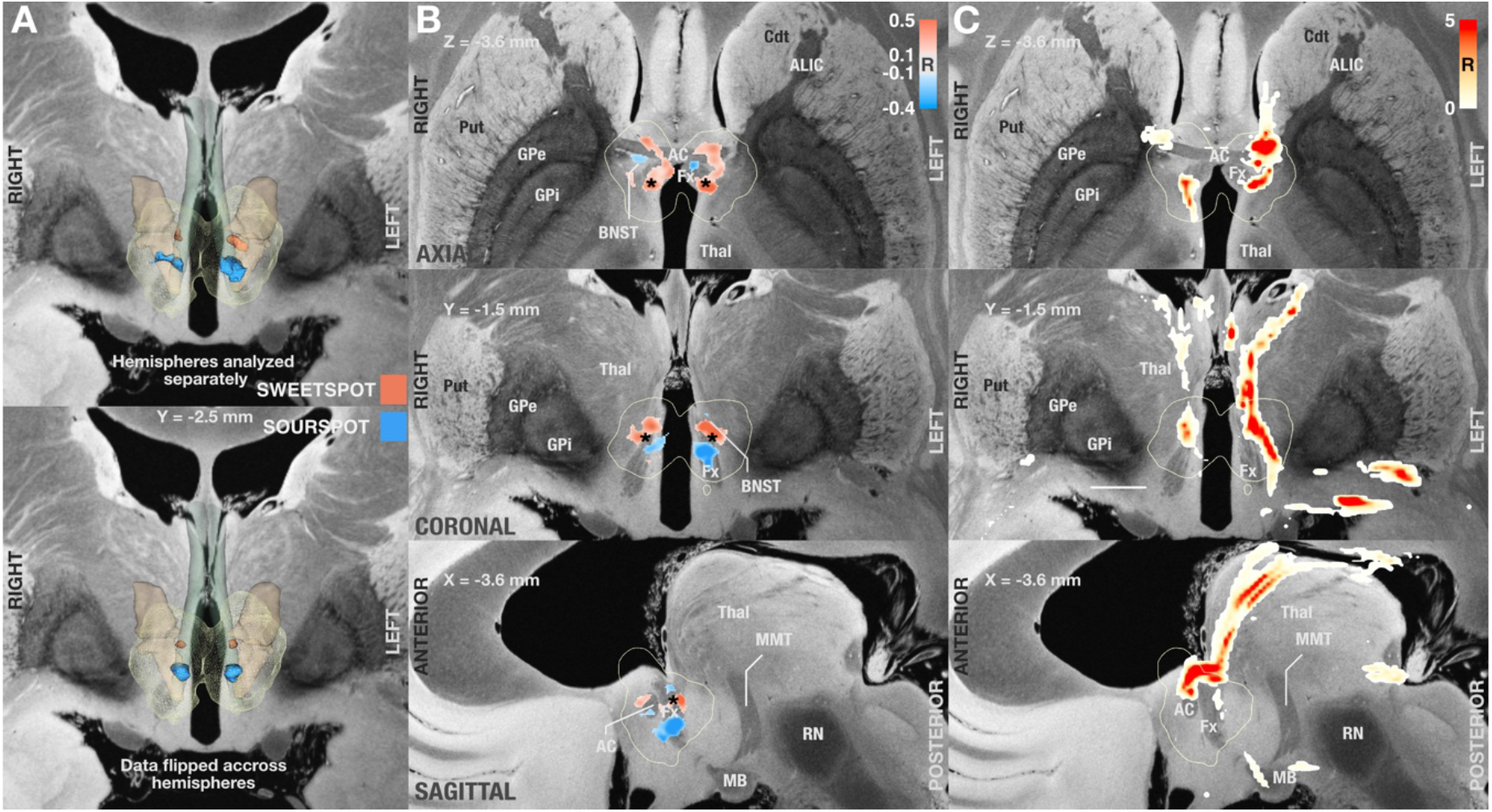
Probabilistic mapping of sweet and sour spots associated with clinical outcome. A) Identified clusters of sweet (red) and sour (blue)-spots in a 3D view, superimposed on slices of a 100-µm, 7T brain scan in MNI 152 space^48^. Since the result was symmetric, on the bottom of the panel, we flipped stimulation volumes across hemispheres to further increase robustness on a voxel-level (effectively doubling the number of electrodes used in each hemisphere). B) Axial, coronal, and sagittal views of sweet and sourspot peak coordinates (also see table S6). Projections of cluster center coordinates are marked by a black asterisk and directly project onto the intersection between fornix and bed nucleus of stria terminalis (BNST, see also Fig. S6). C) Axial, coronal, and sagittal sections showing DBS fiber filtering results obtained from the whole cohort at MNI: X = −3.6, Y = −1.5, and Z = −3.6. Abbr: Put: Putamen, Cdt: Caudate, ALIC: Anterior limb of the internal capsule, AC: Anterior commissure, GPe/i: external / internal pallidum, Thal: thalamus, RN: red nucleus, MB: mamillary bodies, Fx: Fornix. Fornix is shown in blue-green color, informed by the CoBrALab Atlas^47^. Bed nucleus of the stria terminalis shown in light brown color, informed by Neudorfer et al.^39^.

### Distributed Whole-Brain Networks associated with optimal DBS response (DBS network mapping)

Structural connectivity analyses are limited to identification of monosynaptic connections and probabilistic mapping provides insights on a local level. Hence, in an additional analysis, we investigated modulating which functional whole-brain networks was associated with optimal outcomes. To this end, we applied the DBS network mapping method^12,13^ using E-fields as seed regions in a normative connectome calculated from resting-state fMRI scans acquired in 1,000 individuals^42,49^ to identify regions correlated and anti-correlated to functionality of the stimulation volume area (Fig 1C). For each patient, this led to a *fingerprint* of functional connectivity seeding from their respective stimulation sites. Voxel-wise values denoted by these connectivity fingerprints were then correlated with clinical improvements following the approach described by Horn et al^12^. The resulting R-map would show maximal positive values for regions to which connectivity was associated with optimal response, and negative values to regions yielding no clinical benefit (Figure 4). The map was largely symmetric across hemispheres with R-values ranging from −0.45 to 0.43. Optimal response most strongly correlated with connectivity to precuneus, prefrontal regions, cingulate, thalamus, basal ganglia and insula. To validate these results, we again carried out leave-one-out (R = 0.38 at p = 0.006, RMS = 48.69, MAD = 30.99) and several k-fold cross-validation designs (3-fold: R = 0.32 at p = 0.018; 5-fold: R = 0.14 at *p = 0.195*; 7-fold: R = 0.44 at p < 10^16^; 10-fold: R = 0.29 at p = 0.026, Figures 5 and S7). Moreover, repeating the analysis on the training, test and combined cohorts led to highly similar results by visual inspection (Figure 4). To allow a certain degree of reverse inference of these network results^50^, they were spatially compared to maps associated with a total of 1307 terms present in the Neurosynth database (https://neurosynth.org/)^51^. After excluding purely anatomical/functional terms (such as “prefrontal”, “cingulate” or “default”), 7 out of the first 8 cognitive terms related to memory functions or Alzheimer’s Disease, namely: “retrieval”, “memory”, “memory retrieval”, “episodic”, “task”, “demands” and “working memory”. The only outlier term not related to memory, “pain”, ranked at #5. Functional network results and their relationship to cognitive terms are summarized in Figure 4.

**Figure 4:**
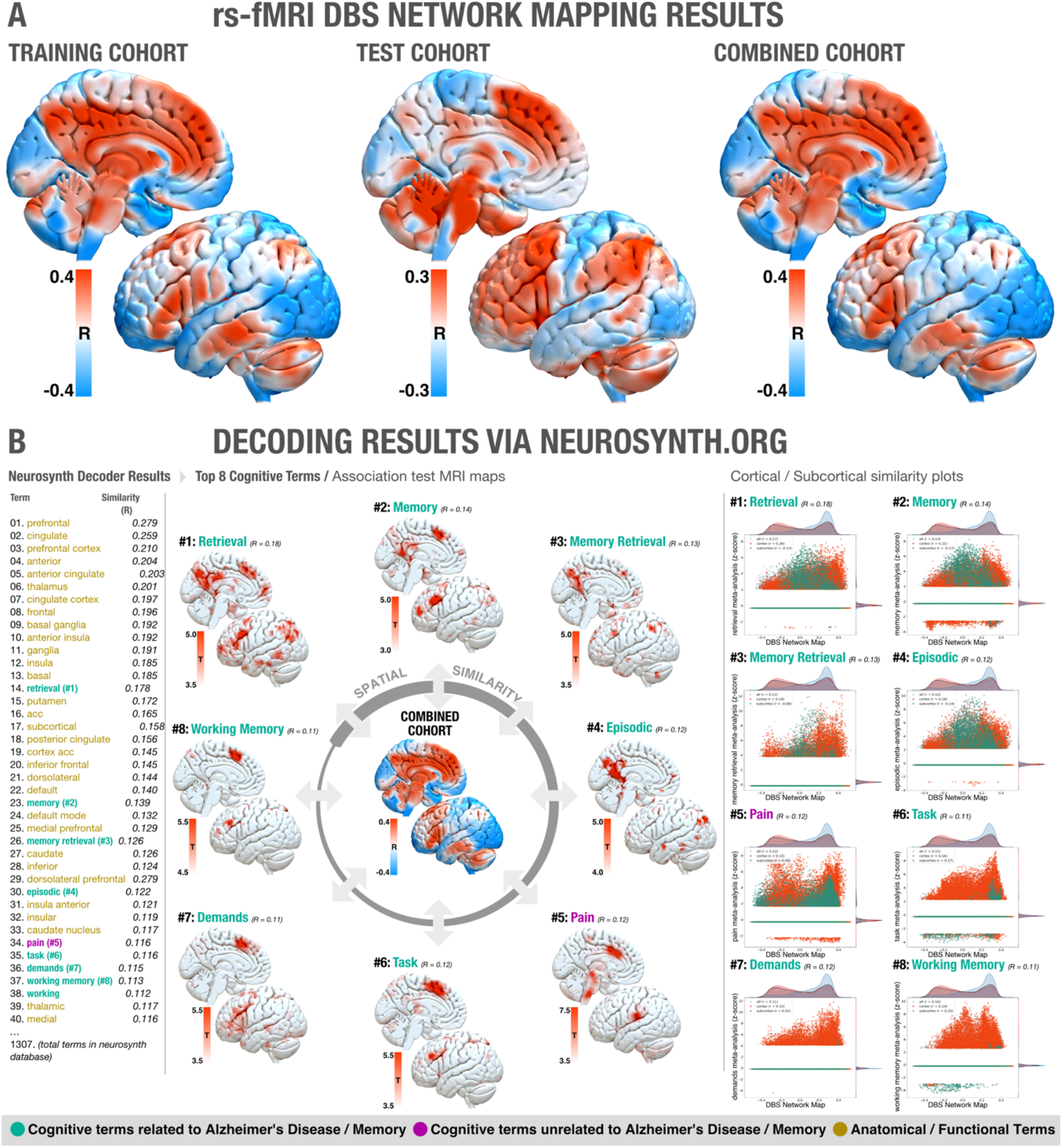
A) Functional networks associated with optimal improvements across training (left), test (middle) and combined (right) cohorts. Brain regions are color-coded by correlations between degree of functional connectivity with DBS electrodes and clinical improvements across the cohorts. Since results were highly symmetric, only the left hemisphere is shown. B) Optimal network associations to Neurosynth database terms, left: highlighted relevant regions for the most similar networks identified; right: similarity plots between same networks and optimal network identified by Network Mapping results (x-axis = specific network meta-analysis, z-score, y-axis = DBS Network Map).

**Figure 5.**
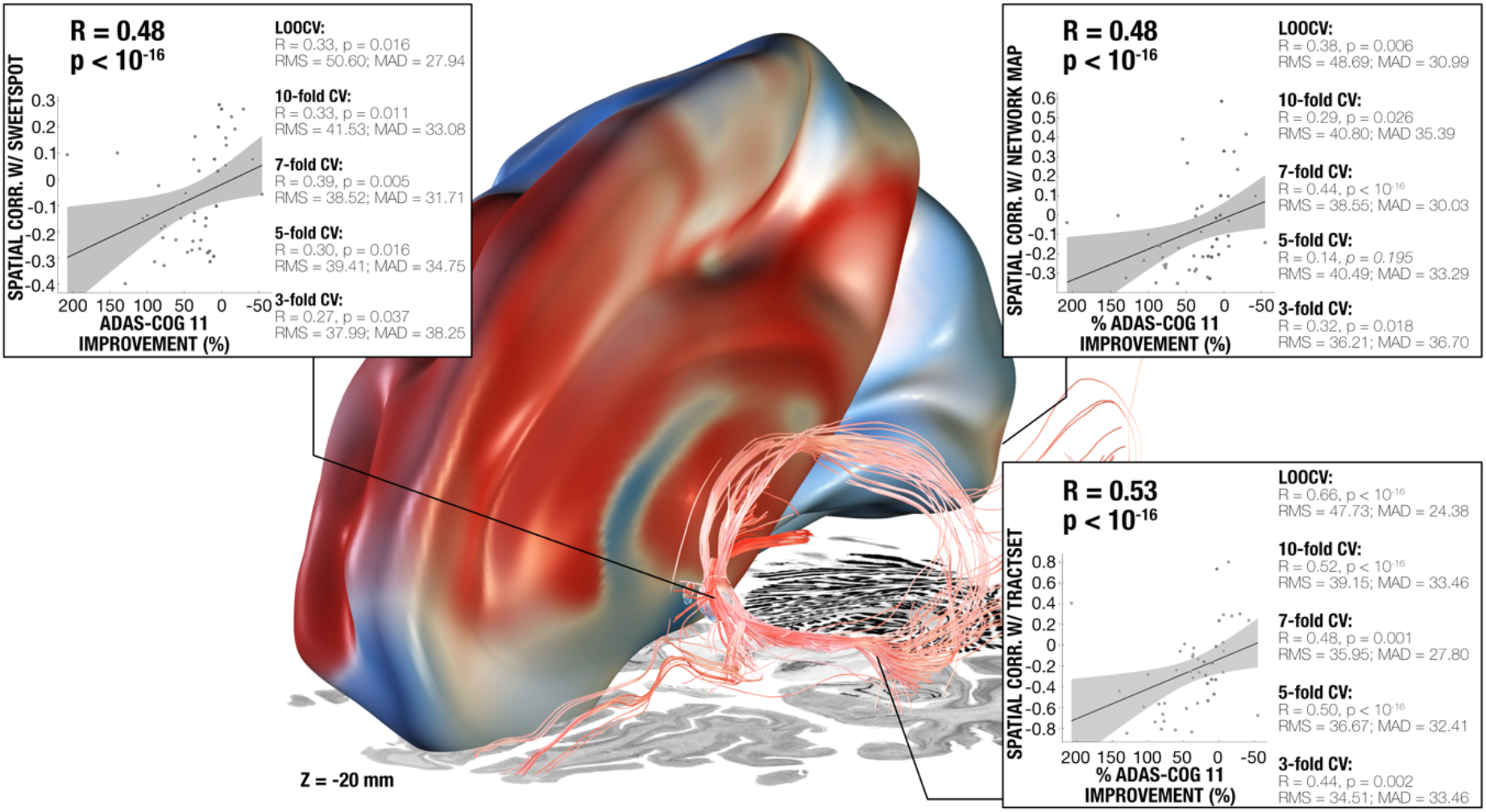
Results summary including the models from DBS fiber filtering, sweetspot mapping and network-mapping. The three levels of analysis were able to explain a similar amount of variance of clinical outcomes when analyzed in a circular nature (see scatterplots; ∼16-19%) and led to significant cross-predictions of clinical outcomes across leave-one-patient-out and multiple k-fold designs. Three level analysis results were superimposed on slices of a brain cytoarchitecture atlas in MNI 152 space^52^. Gray shaded areas represent 95% confidence intervals, see figure S7 for additional metrics on each validation approach.

Figure 5 summarizes the three levels (fiber filtering, sweetspot mapping, and network mapping, see also Figure S7 for in-fold analysis) of analysis and the highly comparable amount of variance explained by each method on a circular (leave-nothing-out) basis, as well as multiple cross-validation designs across the entire cohort. Results (including the same cross-validations) remained highly consistent when repeating all analyses using absolute (instead of relative) improvements on the ADAS-cog 11 scale (Figure S8) and when analyzing the subset of patients enrolled in the ADvance trial (N = 40), in which improvements measured by ADAS-cog 13 were available and applied (Figure S9).

### Effects of age

Prior results had shown differences in clinical improvements related to age groups, where among individuals in the early-on arm during phase 1 (but not in phase 2), participants below the age of 65 worsened on the ADAS-cog13 significantly more than older participants, while those showed improvement^33^. The robustness of models in the present study to successfully cross-estimate clinical improvements across the entire group regardless of age (and regardless of slicing up the data into leave-one-out, 10-, 7-, 5- and 3-fold cross-validation designs) does not a priori confirm such an effect (i.e., the same model seemed to be predictive in both age groups). An alternate reason for age differences could be (potentially atrophy related) systematic shifts in electrode placements. However, as can be seen in figure S10, no apparent difference in electrode placements was observed between the groups, if at all more variability on the z-axis in the young cohort. Furthermore, there was no significant difference in fiber scores obtained across the two age groups (p = 0.790). This does not suggest a systematic shift between groups (such as stimulation in younger participants systematically modulating optimal fiber connections less strongly than in older participants).

### Analysis of Flashback phenomena

In a sub analysis concerning the original hypothesis that led to fx-DBS in AD, we carried out DBS fiber filtering by investigating stimulation settings that did or did not induce flashback-like phenomena during the surgical procedure^30,31^. On a localized level, this effect had been studied before^30,31^, but not using DBS fiber filtering. The sub-cohort in which this information was available included 39 patients in which different DBS parameters were probed, leading to a total of 2054 stimulation volumes, from which 66 resulted in experiential flash-back episodes. In contrast to clinical improvements, flashback-like phenomena were significantly associated with modulation of the posterior limb of the anterior commissure (Figure 6), which interconnects the middle and inferior gyri of the bilateral temporal lobes^53^. Critically, electrical stimulation of these cortical regions has been associated with flashback-like phenomena in multiple historical and contemporary reports^24,54^.

**Figure 6.**
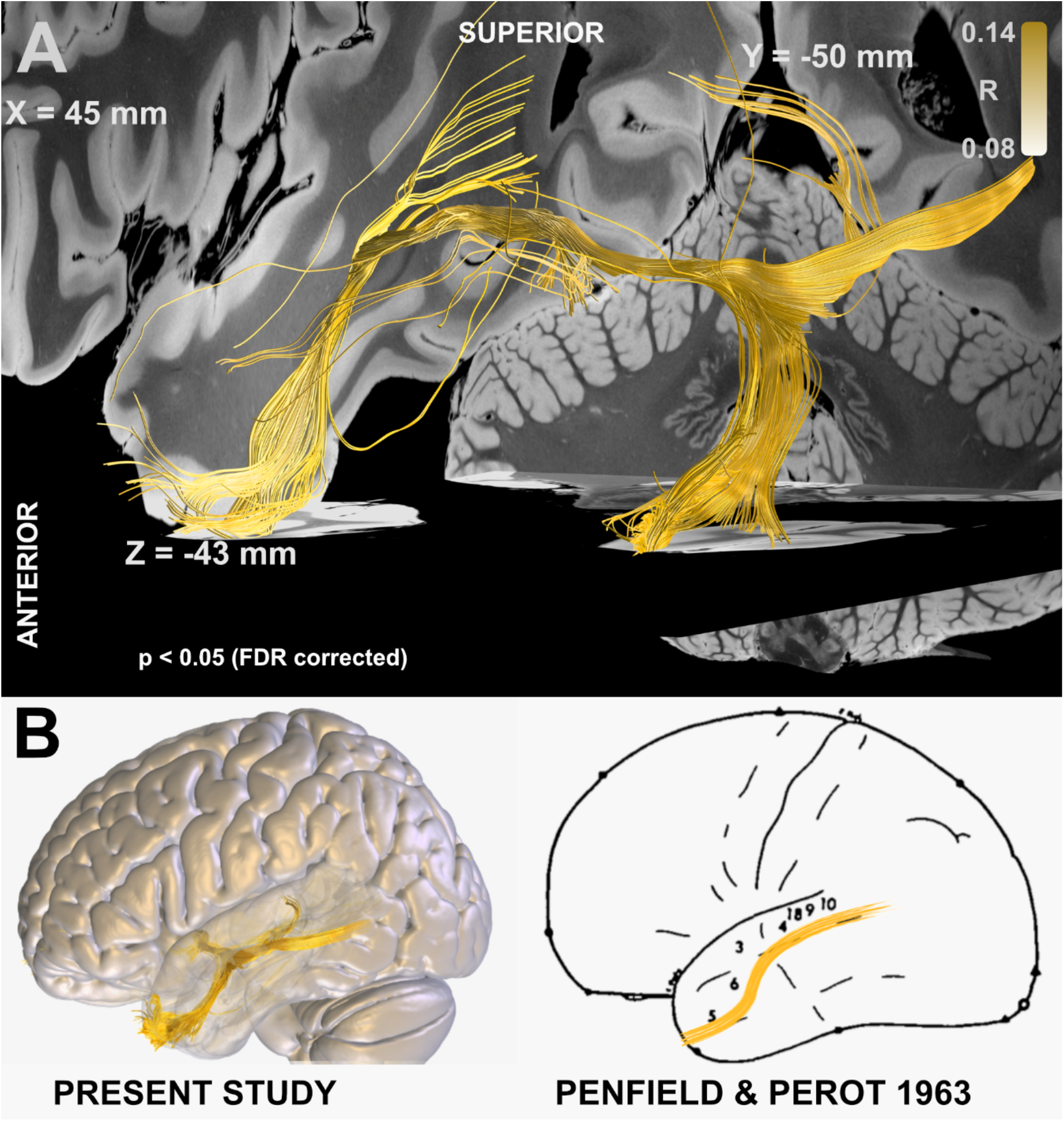
White matter bundle associated with occurrence of flashback-like phenomena. A) Fiber tracts correlated to the presence of flashback-like events. B) Brain surface (lateral view) overlaid with results from A) (left), in comparison to Penfield’s original work on mapping the presence of electrical stimulation-induced “experiential phenomena” in 40 patients suffering from temporal lobe seizures in a total of 1288 reviewed surgical cases covering a large fraction of the cortical mantle (right). Adapted with permission from ^24^.

## Discussion

A three-level post hoc analysis, at the local, structural, and functional connectivity level, was carried out in a cohort of 46 mildly affected AD patients treated with fx-DBS across seven international centers. The results obtained from these analyses provide insights into i) the fiber tracts associated with optimal outcomes, ii) optimal stimulation coordinates (sweetspot maps), and iii) functional whole-brain networks associated with optimal outcomes.

There were many factors that could have led to variability in DBS electrode placements within the fornix region across cases and centers. These factors included decreased fornix volume in AD or its preclinical stage MCI^19^, the complexity of reaching this target using electrodes (transventricular approach)^55^ and the possible variations in placement due to surgeon to surgeon variability. Consequently, electrode localizations varied within the diencephalic region. Considering these variations in DBS lead placement, we sought to examine which white matter pathways were modulated in top-responding but not poor-responding patients. We addressed this question using the DBS fiber-filtering method on tracts defined by an ultra-high-resolution connectome, which was acquired at an isotropic 760 µm resolution, and contains proper definition of fine bundles (such as the stria terminalis) frequently obscured in single-patient scans. Given the historical development of fx-DBS, we hypothesized that fiber tracts associated with optimal response would include memory-relevant connections, specifically the structures of the Papez’ circuit, whose role is crucial in episodic memory^56^ and for which changes have been described as early as in prodromal AD or MCI. This hypothesis was supported by our analyses. Indeed, both fornix and stria terminalis were strongly associated with optimal clinical response. Given the strong implications for clinical practice that our results might have, we cross-validated results on multiple levels, which could demonstrate remarkable consistency of findings throughout subsets of the entire cohort. Based on this white matter model, we were able to estimate a significant amount of variance in clinical outcomes both within the N = 28 training-cohort (leave-one-out and several k-fold designs), and when estimating clinical outcome of patients in the hold-out test-cohort (N = 18) based on the model calculated exclusively from the training cohort. Finally, cross-validations across the N = 46 combined cohort (leave-one-out and several k-fold designs) again showed consistency of findings. Predictive fibers calculated on training and test cohorts alone were remarkably similar, each suggesting a strong involvement of fornix, anterior nuclei of the thalamus and stria terminalis. Interestingly, our analysis yielded a *distinct* set of streamlines when investigating the presence of flashback phenomena reported during postoperative stimulation programming^30,31^. Here, the posterior limb of the anterior commissure emerged as a substrate of modulation. This result supports the main findings from Germann et al.^31^, previously associating stimulation of the anterior commissure with the occurrence of flashbacks, using a different methodological approach.

A seminal historical article by Wilder Penfield and colleagues^24^ associated electrical stimulation of specific sites of the temporal cortex with the occurrence of flashbacks, and this has been recently confirmed by other studies^54^. While the thinner anterior limb of the anterior commissure connects bilateral anterior olfactory nuclei and the primary olfactory cortices, its thicker posterior limb connects the bilateral temporal regions associated with the flashback phenomena reported by cortical stimulation studies^24,54^. Hence, a direct modulation of temporal cortices and/or their network with other structures connected to the anterior commissure might provide a potential reason for the occurrence of flashback phenomena following fornix region DBS.

These tract-level results enhance our understanding of fx-DBS. However, in surgical decision making, defining a focal optimal stimulation coordinate or region could provide additional practical relevance. Hence, we performed a focal analysis to identify a specific sweet spot target associated with clinical improvement. Original surgical coordinates of the active contacts as described for the surgical approach by Ponce et al.^55^ corresponded to an MNI space coordinate^57^ of x = ±7.02 ± 0.68 mm, y = 0.82 ± 1.00 mm and z = −6.43 ± 0.75 mm. In our analysis, we found that cluster centers of positive correlated voxel values instead corresponded to x = ±4.8, y = −0.9 mm and z = −3.6 mm, with a Euclidean distance of 3.87 mm to the original target site. Expressed in functional (AC/PC) coordinates, our target would correspond to a coordinate 5.56 ± 0.88 mm lateral to, −2.87 ± 0.91 mm anterior to and 0.65 ± 1.19 mm below the anterior commissure^57^. Anatomically speaking, the optimal site corresponded to the border between BNST and fornix at a superior (AC level) and posterior portion of the fornix (Figures 3 and S3). Our data suggest that coordinates located more superiorly, and slightly more medial to the current target, may result in better clinical outcomes, a possible explanation might be that the E-field generated when neuromodulating inferior regions of the fornix could be reaching other structures capable of causing side effects, for instance, autonomic responses after hypothalamic nuclei stimulation^32^.

We next applied DBS network mapping using a normative functional connectome to study the relationship between modulating *distributed whole-brain networks* and clinical improvements. In patients with an optimal cognitive response, DBS stimulation sites fell onto a network comprised of regions of the default mode network (especially the precuneus), previously associated with AD pathology^3,4^. Furthermore, the network included premotor cortical sites involved in memory, working memory and retrieval. A common mistake in the fMRI literature is to infer the actual cognitive function from activation (or connection) sites^58,59^. This process, termed reverse-inference, is controversial since activity in most brain regions is often non-specific across cognitive domains. For example, Broca’s area is involved in language processing, but also in other forms of hierarchical processing such as mathematics or music^60^. Hence, (reversely) inferring from an activation in Broca’s area that language is involved would be considered a suboptimal approach^58,59^. To account for this, the creators of the *neurosynth* platform devised a decoding tool facilitating a systematic comparison of network maps with a large amount of meta-analytic maps (N = 1,307 at the time of writing) associated with specific cognitive terms. Each of these maps represents automatic meta-analyses which often rely on a high number of studies – for instance, the map associated with the term *memory* is currently based on 2,744 studies. The decoding tool compares spatial similarity of a given network with all maps in the database, and sorts resulting spatial agreement with term names in descending order. In our case, the functional network most associated with optimal outcome best resembled the maps built from cognitive terms such as “retrieval” or “memory”, hence demonstrating a certain specificity of the identified optimal stimulation network to memory retrieval.

All three levels of analysis (local, tract and network) were highly robust towards multiple cross-validation designs (summarized in Figure 5, in-fold analysis summarized in Figure S7). The findings of this study provide a framework for the neural substrates implicated in successful fx-DBS and offer the potential to refine and guide both surgical targeting and stimulation optimization in Alzheimer’s disease in future trials.

### Limitations

Multiple limitations apply to this work, including the retrospective nature of the study, due to which a detailed focus on specific clinical effects was not possible. For this reason, we considered clinical outcomes as measured by the percent change of the ADAS-cog 11 score but repeated main results for ADAS-cog 13 scores in the subset of patients in which the score was available (figure S9). The retrospective nature of our study also prevented us from analyzing different effects of stimulation frequencies, pulse widths, or stimulation patterns, which would enfold different signals onto the network over time. Instead, the imaging nature of our study analyzes results in static fashion (both on a stimulation volume and network level). Future research is needed to investigate effects of variations in stimulation parameters, such as the ongoing trial to optimize electrical stimulation parameters of fornix-DBS for AD (NCT04856072). Alternatively, neuromodulation delivered through distinct approaches, namely, the ongoing trial on gamma entrainment via sensory stimulus at a 40Hz frequency (NCT04055376) could extend our knowledge on the effect of diverse parameters in brain stimulation for AD.

An inherent limitation of studies as the present one is imaging resolution and resulting inaccuracies of DBS mapping in standardized stereotactic space, which implies co-registration inaccuracies^61^. This inaccuracy could be even more pronounced in AD patients, who characteristically feature structural changes in both white and grey matter, particularly in early onset AD^10,62,63^. To address these issues, a modern DBS imaging pipeline^35^ with advanced concepts such as brain shift correction^64^, multispectral normalization^65^, and phantom validated electrode localizations^66^ was applied. Each processing step was meticulously monitored and corrected, if necessary. In addition, we applied a first-of-its kind manual refinement of normalization warp fields^67^, which was crucial to yield accurate registrations due to large variabilities in patient anatomy. A demonstration of this labor-intensive manual refinement process is visualized in video S1, which shows that upon manual refinements, a good registration accuracy between patient and template fornices was achieved. In this regard, we were not able to find apparent differences in i) electrode placement or ii) fiber-score activations between patients younger than versus older than 65, which suggests other factors might have influenced the clinical outcome in the younger group (Figure S10). As previously reported, possible explanations for the decline in early onset subjects include a more aggressive presentation of the condition, greater brain atrophy and comparably more reduced glucose metabolism in this subgroup of patients^29,60,66,67^.

Another limitation was the combination of randomized and open label outcome data. Due to the exploratory feature of this analysis and aiming at robustness of results, our cohort included patients from different studies, namely a phase I study and the randomized phase II ADvance trial. The inclusion of these two cohorts made it possible to have a large enough sample size to leave a naïve subset of patients to cross-predict our fibertract model.

Moreover, we must emphasize that conclusions about connectivity profiles associated with optimal outcomes were based on normative connectivity data acquired in healthy participants. While this concept has led to meaningful and robust models in other cohorts^12–14,38,71^, conclusions about networks prevalent in the individual DBS patients may not be drawn. However, models describing optimal connectivity based on normative vs. disease-matched vs. patient-specific data were comparable in other diseases, such as Parkinson’s Disease and OCD^38,70^. In the present study, electrodes were placed within the diencephalic region, a region featuring complex neuroanatomical relationships and a multitude of intersecting and delicate fiber bundles. Hence, it was a crucial pre-requisite of the study to use a tractogram that exhibits small fiber bundles in accurate anatomical detail. We used a normative whole-brain connectome calculated from an unprecedentedly high-resolution in-vivo dMRI dataset that was acquired across a total scan time of 18 hours at 760 µm isotropic resolution on specialized MR hardware^70^, as for network mapping, a connectome obtained from rs-fMRI data from 1000 healthy subjects was used to inform regions co-activated with the stimulation volumes of each patient, allowing an identification of circuits that could be involved in clinical changes when modulating the fornix.

### Conclusions

Based on three levels of analysis, our results point towards a *potential optimal stimulation target* for Alzheimer’s Disease treatment with fx-DBS. At a local level, our findings highlight a circumscribed region at the intersection between fornix and bed nucleus of the stria terminalis. We further showed that optimal tract connections to this region contained within the circuit of Papez were important, while flashback phenomena were associated with modulating the posterior limb of the anterior commissure. Finally, our results suggest that modulating specific whole-brain networks is crucial for DBS induced positive effects on cognition. Though our data identified a specific site for stimulation, we would like to emphasize that the use of indirect coordinate systems for DBS targeting is not suitable for DBS to the fornix region in patients with atrophy in the same region. Direct imaging and fiber-tracking results will be important to determine accurate targeting in this region.

## Methods

### Patient cohort and Imaging

We conducted a secondary post-hoc analysis of data from a sample of 46 patients (mean age: 67 ± 7.9 years, 22 females), with a clinical diagnosis of mild probable AD that underwent bilateral DBS to the fornix at seven international centers included in the ADvance trial (NCT01608061)^33^ and the Toronto-based pilot trial (NCT00658125)^25^. While the ADvance trial included 42 patients, imaging data was only available for 40 patients ^31,32^ (also see figure S1). Patients were diagnosed by standardized criteria after expert examination rated with 0.5 or 1 on the Clinical Dementia Rating scale (CDR) and scored 12-24 on the Alzheimer’s Disease Assessment Scale 11 (ADAS-cog)^72^, further inclusion and exclusion criteria for the trials can be found in tables S1 and S2, patients received monopolar stimulation at a frequency of 130 Hertz with a 90 millisecond pulse width for 12 months without adjustment. Patients included in the ADvance trial were evaluated using the ADAS-cog 13 scale but for remaining patients, improvements along ADAS-cog 11 was available. Hence, for consistency across the entire cohort, two tasks were excluded from this scale (number cancellation task and delayed free recall task)^73^, and only tasks included in ADAS-cog 11^74^ were included for analysis. We repeated main analyses using ADAS-cog 13 in the subset of patients in which the score was available. Patients underwent surgery targeting the descending columns of the fornix using quadripolar electrodes (Medtronic 3387, Medtronic, Minneapolis, MN). T^1^- and T^2^-weighted volumetric pre- and postoperative scans obtained at 1.5 T across seven sites were used. Intra- and post-operative test stimulation observations, and individual stimulation parameters including electrode contact, stimulation amplitude, frequency, and pulse width were included. The additional post hoc data analysis carried out in the present study was approved by the ethics board of Charité – Universitätsmedizin Berlin (master vote EA2/186/18). The clinical outcome of all patients was evaluated using the ADAS-cog 11^74^ measured before and one year after the onset of stimulation. Exclusively for means of visualization, participants were classified according to their ADAS-cog 11 outcome as poor responders (increase of 21%), middle responders (0-to-20.99% increase), top responders (decrease in ADAS-cog 11 score). This classification was not used for statistical analyses, which were carried out on the continuous outcome variable (percentage outcome on ADAS-cog 11 one year after stimulation onset) and the discrete variable TEMPau score^75^, used to estimate flashback episode intensities.

### DBS electrode localization and stimulation volume (E-field) estimation

Image pre-processing, electrode localization and estimation of stimulation volume were carried out using default parameters in Lead-DBS^35,64^ (www.lead-dbs.org). Briefly, post-operative MRI scans were linearly co-registered to preoperative T1 images using Advanced Normalization Tools^65^ (ANTs; http://stnava.github.io/ANTs/). Subcortical refinement was applied to correct for brain shift. Co-registered images were then normalized into ICBM 2009b Nonlinear Asymmetric (“MNI”) template space using the SyN approach implemented in ANTs, with an additional subcortical refinement stage to attain a most precise subcortical alignment between patient and template space (“Effective: Low Variance + subcortical refinement” preset). While this method has been shown to yield the best performance for subcortical image registrations^76^, the substantial atrophy in this particular population resulted in suboptimal automatic registration results. For the present study, this was crucial, since in the field of DBS, electrode displacements of a few millimeters will lead to substantially different effects^35,37^. To account for this, we applied a novel method, termed WarpDrive^67^, to manually refine registrations into template space (see video S1). Briefly, WarpDrive provides a graphical interface allowing precise alignment of source and target landmarks by directly visualizing the normalized images, together with the template and atlases in MNI space (the software is openly available here: https://github.com/netstim/SlicerNetstim). WarpDrive allows the user to manually correct misalignments from the standard normalization and recomputes a refined deformation field in real time. DBS electrodes were pre-localized using the TRAC/CORE algorithm^64^ and manually refined if necessary. Stimulation volumes were estimated using the finite element method (FEM) within the adapted FieldTrip / SimBio pipeline^77^ (https://www.mrt.uni-jena.de/simbio/; http://fieldtriptoolbox.org/) implemented in Lead-DBS^35^. In brief, a volume conductor model was constructed based on a four-compartment mesh that included gray and white matter, electrode contacts and insulating parts. Gray matter structures were based on an atlas of the human hypothalamic region^39^. The electric field (E-field) distribution was then estimated by solving the static formulation of the Laplace equation on a discretized domain represented by the tetrahedral four-compartment mesh. For the purpose of this article, we occasionally use *E-field* as shorthand for the voxelized magnitude of the electric field vector. The stimulation volumes were defined as *thresholded* copies of the E-field magnitude following the approach in^45^.

### Modeling considerations

Estimated after Pakkenberg and colleagues^78^, each cubic millimeter of cortex is filled with ∼170,000 neurons; for axonal numbers, each fiber bundle in a standard neuroimaging analysis represents 10^3^-10^5^ tightly packed axons^79^. Many DBS studies aimed at modeling discretized and realistic axonal cable models, in the past^39–41^. However, given these sheer numbers of axons involved, here, we chose to assume probabilistic axonal populations in each brain voxel to be represented by each fiber tract, instead of modeling representative single axons. While single axons fire in an all-or-nothing fashion, activation/modulation profiles of axonal populations within a voxel may be represented in probabilistic fashion, which would be dependent on the applied voltage^80–82^. In other words, on a population level, the “degree” of activation will be stronger under higher voltages applied, i.e., closer to the electrodes. Crucially, there is large amount of uncertainty about this exact relationship between voltage and population-level axonal firing that needs patient-specific calibration even when applying more realistic biophysical models^83^. To account for this uncertainty, we applied Spearman’s rank correlations to our fiber filtering and optimal stimulation site mapping models (Figure 1). We propose that this simple approach could be advantageous, since it would show maximal correlations for any type of monotonically increasing dose-effect function. In other words, the concept could be robust toward the exact relationship (be it e.g., linear, cubic, or logistic) between amplitude and axonal modulation.

For each of the models, the stimulation volume of each patient was considered the core of the analysis; for fiber filtering, streamlines from a normative structural connectome that traversed the volumes were considered for further steps; for sweetspot analysis, areas of interest were determined based on voxels occupied by stimulation volumes of the patients; finally, for network mapping, functionally connected areas to the stimulation volume of each patient were obtained from a functional normative connectome. Details for each method are specified in the following sections.

### DBS fiber filtering

#### Model definition (Figure 1A)

Whole brain structural connectivity profiles seeding from bilateral E-fields were calculated using a state-of-the-art multi-shell diffusion-weighted imaging dataset acquired across 18 scanning hours of a single individual at 760 µm isotropic resolution^41^ using the generalized q-sampling approach (default parameters) and whole-brain tracking (default parameters) as implemented in DSI studio^84^. The patients were distributed into two cohorts: Training (N = 28) and Test (N = 18). For each subject of the training cohort, fibers traversing each voxel of the E-field were selected from the 5 million tracts in the normative connectome and projected to a voxelized volume in MNI space. Each of these fibers were weighted according to the E-field magnitude at each voxel, considering only fibers that traversed > 20% of stimulation volumes with an E-field magnitude > 0.36 V/mm. Each fiber was then appointed an R-value dependent on the Spearman correlation between its weighting and the respective clinical outcome scores across the group, i.e., a high R-value indicates that the modulation of that tract is associated with clinical improvement. Given the mass-univariate nature of this approach (and subsequent alpha-error accumulations), the resulting correlation coefficients were not considered significant, but were rather used to discriminate and visualize a specific set of bundles that was later validated by estimating clinical outcome in out of sample data (Figure 1, table S5).

#### Estimating outcomes using the model

Assuming a patient would most likely show superior clinical benefit if their E-field modulated more fibers with high positive R-values and less fibers with negative scores, we measured the spatial Spearman’s rank correlation profile of the (unseen) E-field superimposed to the tract model. To illustrate by two examples: If an E-field peaked at sites coinciding with tracts with high positive R-values and showed low amplitudes at sites filled by tracts with low R-values, this would lead to a high Fiber-Score for that particular E-field.

#### Cross-validation and testing

We first estimated our model by defining a “Training cohort” including 60% of the participants in a random fashion, and filtering fibers with positive R-values across this group; the remaining 40% of the participants (“Test cohort”) were left to validate the predictive utility of the model. The *Training cohort* was used to estimate an *optimal connectivity model*. In an initial training stage (using only data from the training cohort), model parameters were still manually tuned using the graphical user interface created for Fiber Filtering within Lead-DBS. Aims were to obtain a set of fibers that was i) robust for cross-validations and ii) not robust for permuted improvement values. The latter point was crucial, since specific parameter settings exist that would result in a set of tracts that were simply connected with the average group of electrodes. In such settings, permuting improvement values across the cohort would not largely alter results. After several iterations, settings were obtained (table S5) that fulfilled both criteria and showed robust cross-validation results (leave one out and multiple k-fold [k=3,5,7,10] designs). Then, model parameters were kept fixed and, only then was the model used to cross-predict outcomes of patients in the hold-out *test cohort*.

### Optimal Stimulation Sites (Sweetspot Analysis)

#### Model definition (Figure 1B)

Using the E-fields calculated for each patient, an approach to define optimal stimulation sites was applied^12^. E-fields represent the first derivative of the estimated voltage distribution applied to voxels in space and their vector magnitudes are hence stronger in proximity of active electrode contacts with a rapid decay over distance. Since not all voxels were covered by the same number of E-fields, the area of interest was restricted to voxels that were at least covered by 20% of E-fields with a vector magnitude above 200 V/m, which is a common approximate assumed to activate axons in the field of DBS^44^. For each voxel covered by the group of E-fields across the cohort in MNI space, E-field vector magnitudes across patients were Spearman rank correlated with clinical outcomes. The resulting sweetspot maps would peak at voxels in which stronger E-fields were associated with better treatment responses. The map would have negative values for voxels with the opposite relationship.

#### Estimating outcomes using the model

Multiplying each voxel of a single E-field with the resulting sweetspot map and calculating the average across voxels led to estimates of how a specific E-field would perform (i.e., estimates of clinical outcomes following DBS). If the E-field peaked at similar locations as the sweetspot map, a high estimate would result. If it would peak at a valley of the map, low or even negative estimates would result. The values of these maps where analyzed using Multi-image Analysis GUI) software^85^ (http://ric.uthscsa.edu/mango/) to estimate the peak and center location of clusters in both positive (sweetspot) and negative (sourspot) correlated voxels, this analysis was repeated in E-fields mirrored to opposite hemispheres to obtain a more robust observation of peak voxels. Again, cross-validation of the model was carried out by means of a Leave-one-out and several k-fold [k=3,5,7,10] designs.

### DBS Network Mapping

#### Model definition (Figure 1C)

In a third approach, we calculated whole-brain functional connectivity estimates seeding from E-fields using a normative connectome that was calculated from rs-fMRI scans acquired in 1,000 healthy participants, providing a map of coupling brain regions based on their blood-oxygen-level-dependent (BOLD) signal^42,49^, following the approach developed by Horn et al^12^. This method, termed DBS network mapping, allowed to investigate functional connectivity profiles of a specific pair of DBS electrodes. We refer to the maps resulting from an estimation of correlated “active” brain regions seeded from each stimulation volume using normative data as *connectivity fingerprints*^14^. Similar to the sweetspot and fiber filtering models, (voxel-wise) correlations between Fisher-z-scored connectivity strengths and clinical improvements were calculated, which led to R-map models of *optimal connectivity*.

### Estimating outcomes using the model

In direct parallel to the other two approaches, spatial similarities between single connectivity fingerprints and R-map models were calculated using voxel-wise spatial correlations. This led to positive high correlation values for cases in which fingerprints graphically matched the *(optimal) connectivity profile* represented by the R-map model – and lower or even negative values for the opposite cases. The R-model obtained by combining all the single connectivity fingerprints was cross-validated using a leave-one-out and multiple k-fold [k=3,5,7,10] designs, and quantitatively and interactively compared to the *Neurosynth* database (neurosynth.org) to allow comparison of the identified map to functional networks previously reported by other studies.

Analyses on the three levels (fiber filtering, sweetspot mapping and functional network mapping) were repeated using absolute (instead of relative) improvements of ADAS-cog 11 following DBS (Figure S8), as well as improvements measured by ADAS-cog 13 scores (Figure S9). In the latter, only the subset of patients from the ADvance trial were included (since in other patients, ADAS-cog 13 improvements were not available).

### Analysis of Flashback-like Phenomena

During the surgical intervention of a subset of the cohort, 39 patients aged 67.7 ± 8 years old, 19 females (participants from ADvance fx-DBS trial^33^, NCT01608061), it was tested whether flashback phenomena could be induced^30,31^ by means of stimulation with increasing voltages (1-10) at multiple contacts (0-3), eliciting at least one memory flashback in 18 (8 females) of these patients. This resulted in a total of 2054 stimulation volume probes, 66 of which evoked a flashback-like episode. We investigated the presence of streamlines correlated to these stimulation volumes in the same fashion as we did our whole dataset using the Fiber Filtering Tool (Figure 1A).

## Data Availability

Anonymized derivatives of stimulation data used for the described analyses are openly available on OSF (https://osf.io/bckuf). The resulting tract atlas, sweet spot and fMRI network pattern are openly available within Lead-DBS software (www.lead-dbs.org).

## Code availability

All code used to analyze the dataset is openly available within Lead-DBS/-Connectome software (https://github.com/leaddbs/leaddbs). Code to reproduce figures is openly available on OSF (https://osf.io/bckuf).

## Acknowledgements

C.N. was supported by the German Research Foundation (Deutsche Forschungsgemeinschaft, DFG NE 2276/1-1). K.F. received grants and personal fees from Medtronic and Boston Scientific, grants from Abbott/St. Jude, and Functional Neuromodulation outside the submitted work. D.W. received grants from Functional Neuromodulation during conduct of this study, grants and personal fees from Avid/Lily, and Merck, personal fees from Jannsen, GE Healthcare, Biogen and Neuronix outside the submitted work. S.S. receives personal fees from Elsai, Lilly, Roche Novartis and Biogen outside the submitted work. M.S. received personal fees from Allergan, Biogen, Roche-Genentech, Cortexyme, Bracket, Sanofi, and other type of support from Brain Health Inc and uMethod Health outside of the submitted work. C.L. received grants from Functional Neuromodulation Inc. during conduct of this study, from Avanir and Eli Lily and NFL Benefits Office outside of the submitted work. M.O. received grants from NIH, Tourette Association of America Grant, Parkinson’s Alliance, Smallwood Foundation, and personal fees from Parkinson’s Foundation Medical Director, Books4Patients, American Academy of Neurology, Peerview, WebMD/Medscape, Mededicus, Movement Disorders Society, Taylor and Francis, Demos, Robert Rose and non-financial support from Medtronic outside of the submitted work. A.L. received grants from Medtronic and Functional Neuromodulation during conduct of this study, personal fees from Medtronic, St. Jude, Boston Scientific, and Functional Neuromodulation outside of submitted work. A.L. disclosed having a patent “US Patent 8,346,365. Lozano AM. Cognitive function within a human brain. 2013” licensed to Functional Neuromodulation. A.H. was supported by the German Research Foundation (Deutsche Forschungsgemeinschaft, 424778381 – TRR 295), Deutsches Zentrum für Luft-und Raumfahrt (DynaSti grant within the EU Joint Programme Neurodegenerative Disease Research, JPND), the National Institutes of Health (2R01 MH113929) as well as the New Venture Fund (FFOR Seed Grant). ADvance was supported by the National Institute on Aging (R01AG042165) and Functional Neuromodulation Ltd., the sponsor of the study. Other co-authors report no conflicts of interest.

## The ADvance Study team included

### Functional Neuromodulation

Todd Langevin, Lisa Fosdick, Kristen Drake, Donald E. Reymers, Robyn Moxon, Dan O’Connell, Vince Owens, Cara Pendergrass, Susan Klees, Steven D. Targum. and the seven participating clinical trial sites:

### Chair’s Office at Johns Hopkins University and University of Toronto

Constantine G. Lyketsos, MD, MHS, Co-PI, Elizabeth Plank Althouse Professor and Chair of Psychiatry and Behavioral Sciences at Johns Hopkins Bayview; Andres M. Lozano, MD, PhD, FRCSC, FACS, Co-PI, Professor, and Chair of Neurosurgery, Tasker Chair of Functional Neurosurgery; Gwenn Smith, PhD, Imaging Core Director, Richman Family Professor of Psychiatry and Behavioral Sciences, Johns Hopkins University; Cynthia Munro, PhD, Neuropsychologist, Associate Professor of Psychiatry and Behavioral Sciences, Johns Hopkins University; Esther Oh, MD, Medical Monitor, Assistant Professor of Geriatric Medicine, Johns Hopkins University; Jeannie Sheppard Leoutsakos, PhD, Data Core Leader, Assistant Professor of Psychiatry and Behavioral Sciences, Johns Hopkins University.

### Clinical Trial Sites

*Banner Alzheimer’s Institute, Phoenix*: Anna Burke, MD, Geriatric Psychiatrist, Dementia Specialist; Francisco A. Ponce, MD, Associate Professor of Neurosurgery, Director Barrow Center for Neuromodulation.

*Banner Sun Health Research Institute, Sun City*: Marwan Sabbagh, MD, Director, Banner Sun Health Research Institute; Francisco A. Ponce, MD, Associate Professor of Neurosurgery, Director Barrow Center for Neuromodulation.

*Brown University, Rhode Island Hospital, Butler Hospital*: Stephen Salloway, MD/MS Professor of Neurology, Director of Neurology and Memory and Aging Program; Rees Cosgrove, MD/PhD, Chair of Neurosurgery; Wael Asaad, MD/PhD, Assistant Professor of Neurosurgery.

*Johns Hopkins University School of Medicine, Baltimore MD*: Paul Rosenberg, MD, Associate Professor, Associate Director, Memory and Alzheimer’s Treatment Center; William S. Anderson, MD, PhD, Associate Professor of Neurosurgery, Zoltan Mari, M. D, Associate Professor of Neurology, Ned Sacktor, MD, Professor of Neurology.

*University of Florida – Gainesville*: Michael S. Okun, MD, Professor of Neurology, Co-Director of the Center for Movement Disorders and Neurorestoration; Kelly D. Foote, MD, Professor of Neurosurgery, Co-Director for Center for Movement Disorders and Neurorestoration.

*University of Pennsylvania*: David A. Wolk, MD, Associate Professor of Neurology, Assistant Director Penn Memory Center; Gordon Baltuch, MD/PhD, Professor of Neurosurgery, Director Center for Functional and Neurorestorative Neurosurgery.

*University of Toronto/Toronto Western Hospital*: Andres M. Lozano, MD, PhD, FRCSC, FACS, Professor of Neurosurgery, Tasker Chair of Functional Neurosurgery; David F. Tang-Wai, MDCM FRCPC, Associate Professor of Neurology.

## Author Contributions

ASRI and AH designed the study, analyzed the data, and wrote the manuscript. AB, GJBE, AL, WD, BS, LA, KDF, RA, JG, PBR, DFTW, DAW, ADB, SS, MNS, MMC, GSS, CGL, MSO, WSA, ZM, FAP, AML collected the data and critically revised the manuscript. SO, CN, KB, NL and NR analyzed the data and critically revised the manuscript, KB performed in-fold analysis.

## References

1. Winston, W. Economic Burden of Alzheimer Disease and Managed Care Considerations. Am J Manag Care 26, S177–S183 (2020).

2. Hyman, B. T. et al. National Institute on Aging-Alzheimer’s Association guidelines for the neuropathologic assessment of Alzheimer’s disease. Alzheimers Dement 8, 1–13 (2012).

3. Greicius, M. D., Srivastava, G., Reiss, A. L. & Menon, V. Default-mode network activity distinguishes Alzheimer’s disease from healthy aging: Evidence from functional MRI. Proceedings of the National Academy of Sciences 101, 4637–4642 (2004).

4. Mevel, K., Chételat, G., Eustache, F. & Desgranges, B. The default mode network in healthy aging and Alzheimer’s disease. Int J Alzheimers Dis 2011, 535816 (2011).

5. Canter, R. G., Penney, J. & Tsai, L.-H. The road to restoring neural circuits for the treatment of Alzheimer’s disease. Nature 539, 187–196 (2016).

6. Thal, D. R., Rüb, U., Orantes, M. & Braak, H. Phases of A beta-deposition in the human brain and its relevance for the development of AD. Neurology 58, 1791–1800 (2002).

7. Braak, H. & Braak, E. Neuropathological stageing of Alzheimer-related changes. Acta Neuropathol 82, 239–259 (1991).

8. Jeong, J. EEG dynamics in patients with Alzheimer’s disease. Clin Neurophysiol 115, 1490–1505 (2004).

9. Jacobs, H. I. L., Radua, J., Lückmann, H. C. & Sack, A. T. Jacobs, H. I., Radua, J., Lückmann, H. C., & Sack, A. T. Meta-analysis of functional network alterations in Alzheimer’s disease: toward a network biomarker. Neuroscience and biobehavioral reviews 37, 753–765 (2013).

10. Kitamura, S. et al. Longitudinal white matter changes in Alzheimer’s disease: A tractography-based analysis study. Brain Research 1515, 12–18 (2013).

11. Lozano, A. M. & Lipsman, N. Probing and Regulating Dysfunctional Circuits Using Deep Brain Stimulation. Neuron 77, 406–424 (2013).

12. Horn, A. et al. Connectivity Predicts deep brain stimulation outcome in Parkinson disease. Ann Neurol 82, 67–78 (2017).

13. Al-Fatly, B. et al. Connectivity profile of thalamic deep brain stimulation to effectively treat essential tremor. Brain 18, 130 (2019).

14. Li, N. et al. A unified connectomic target for deep brain stimulation in obsessive-compulsive disorder. Nature Communications 11, 3364 (2020).

15. Horn, A. et al. Differential Deep Brain Stimulation Sites and Networks for Cervical vs. Generalized Dystonia. 2021.07.28.21261289 (2021) doi:10.1101/2021.07.28.21261289.

16. Iaccarino, H. F. et al. Gamma frequency entrainment attenuates amyloid load and modifies microglia. Nature 540, 230–235 (2016).

17. Martorell, A. J. et al. Multi-sensory Gamma Stimulation Ameliorates Alzheimer’s-Associated Pathology and Improves Cognition. Cell 177, 256-271.e22 (2019).

18. Aldehri, M., Temel, Y., Alnaami, I., Jahanshahi, A. & Hescham, S. Deep brain stimulation for Alzheimer’s Disease: An update. Surg Neurol Int 9, 58 (2018).

19. Fletcher, E. et al. Loss of Fornix White Matter Volume as a Predictor of Cognitive Impairment in Cognitively Normal Elderly Individuals. JAMA Neurol 70, 1389–1395 (2013).

20. Mielke, M. M. et al. Fornix integrity and hippocampal volume predict memory decline and progression to Alzheimer’s disease. Alzheimer’s & Dementia 8, 105–113 (2012).

21. Oishi, K. & Lyketsos, C. G. Alzheimer’s disease and the fornix. Frontiers in Aging Neuroscience 6, 241 (2014).

22. Nowrangi, M. A. & Rosenberg, P. B. The Fornix in Mild Cognitive Impairment and Alzheimer’s Disease. Frontiers in Aging Neuroscience 7, 1 (2015).

23. Hamani, C. et al. Memory enhancement induced by hypothalamic/fornix deep brain stimulation. Ann Neurol. 63, 119–123 (2008).

24. Penfield, W. & Perot, P. The Brain’s Record of Auditory and Visual Experience: A Final Summary and Discussion. Brain 86, 595–696 (1963).

25. Laxton, A. W. et al. A phase I trial of deep brain stimulation of memory circuits in Alzheimer&s disease. Ann Neurol 68, 521–534 (2010).

26. Scharre, D. et al. Deep Brain Stimulation of Frontal Lobe Networks to Treat Alzheimer’s Disease. Journal of Alzheimer’s Disease 62, 1–13 (2018).

27. Baldermann, J. C. et al. Neuroanatomical Characteristics Associated With Response to Deep Brain Stimulation of the Nucleus Basalis of Meynert for Alzheimer&s Disease. Neuromodulation: Technology at the Neural Interface 26, 2411 (2017).

28. Bittlinger, M. & Müller, S. Opening the debate on deep brain stimulation for ALzheimer’s disease - a critical evaluation of rationale, shortcomings, and ethical justification. BMC Med Ethics 19, (2018).

29. Lozano, A. M. et al. A Phase II Study of Fornix Deep Brain Stimulation in Mild Alzheimer&s Disease. - PubMed - NCBI. JAD 54, 777–787 (2016).

30. Deeb, W. et al. Fornix-Region Deep Brain Stimulation–Induced Memory Flashbacks in Alzheimer’s Disease. New England Journal of Medicine 381, 783–785 (2019).

31. Germann, J. et al. Brain structures and networks responsible for stimulation-induced memory flashbacks during forniceal deep brain stimulation for Alzheimer’s disease. Alzheimer’s & Dementia 17, 777–787 (2021).

32. Neudorfer, C. et al. Mapping autonomic, mood and cognitive effects of hypothalamic region deep brain stimulation | Brain | Oxford Academic. Brain 144, 2837–2851 (2021).

33. Leoutsakos, J.-M. S. et al. Deep Brain Stimulation Targeting the Fornix for Mild Alzheimer Dementia (the ADvance Trial): A Two Year Follow-up Including Results of Delayed Activation. Journal of Alzheimer’s Disease 64, 597–606 (2018).

34. Baldermann, J. C. et al. Connectomic deep brain stimulation for obsessive-compulsive disorder. Biological Psychiatry (2021) doi:10.1016/j.biopsych.2021.07.010.

35. Horn, A. et al. Lead-DBS v2: Towards a comprehensive pipeline for deep brain stimulation imaging. NeuroImage 184, 293–316 (2019).

36. Treu, S. et al. Deep brain stimulation: Imaging on a group level. NeuroImage 219, 117018 (2020).

37. Horn, A. The impact of modern-day neuroimaging on the field of deep brain stimulation. Current Opinion in Neurology 32, 511–520 (2019).

38. Baldermann, J. C. et al. Connectivity Profile Predictive of Effective Deep Brain Stimulation in Obsessive-Compulsive Disorder. Biol. Psychiatry 85, 735–743 (2019).

39. Neudorfer, C. et al. A high-resolution in vivo magnetic resonance imaging atlas of the human hypothalamic region. Sci Data 7, 305 (2020).

40. Dembek, T. A. et al. Sweetspot Mapping in Deep Brain Stimulation: Strengths and Limitations of Current Approaches. Neuromodulation: Technology at the Neural Interface (2021) doi:https://doi.org/10.1111/ner.13356.

41. Wang, F. et al. In vivo human whole-brain Connectom diffusion MRI dataset at 760 µm isotropic resolution. Scientific Data 8, 122 (2021).

42. Holmes, A. J. et al. Brain Genomics Superstruct Project initial data release with structural, functional, and behavioral measures. Sci. Data 2, 1–16 (2015).

43. Choi, K. S., Riva-Posse, P., Gross, R. E. & Mayberg, H. S. Mapping the “Depression Switch” During Intraoperative Testing of Subcallosal Cingulate Deep Brain Stimulation. JAMA Neurology 72, 1252–1260 (2015).

44. Irmen, F. et al. Left prefrontal impact links subthalamic stimulation with depressive symptoms. Ann Neurol. 87, 962–975 (2020).

45. Astrom, M., Diczfalusy, E., Martens, H. & Wardell, K. Relationship between neural activation and electric field distribution during deep brain stimulation. IEEE Trans Biomed Eng 62, 664–672 (2015).

46. Poldrack, R. A., Huckins, G. & Varoquaux, G. Establishment of Best Practices for Evidence for Prediction: A Review. JAMA Psychiatry 77, 534–540 (2020).

47. Amaral, R. S. C. et al. Manual segmentation of the fornix, fimbria, and alveus on high-resolution 3T MRI: Application via fully-automated mapping of the human memory circuit white and grey matter in healthy and pathological aging. Neuroimage 170, 132–150 (2018).

48. Edlow, B. L. et al. 7 Tesla MRI of the ex vivo human brain at 100 micron resolution. Sci. Data 6, 244 (2019).

49. Yeo, B. T. T. et al. The organization of the human cerebral cortex estimated by intrinsic functional connectivity. J Neurophysiol 106, 1125–1165 (2011).

50. Rubin, T. N. et al. Decoding brain activity using a large-scale probabilistic functional-anatomical atlas of human cognition. PLOS Computational Biology 13, e1005649 (2017).

51. Yarkoni, T., Poldrack, R. A., Van Essen, D. C. & Wager, T. D. Cognitive neuroscience 2.0: building a cumulative science of human brain function. Trends in Cognitive Sciences 14, 489–496 (2010).

52. Amunts, K., Mohlberg, H., Bludau, S. & Zilles, K. Julich-Brain: A 3D probabilistic atlas of the human brain’s cytoarchitecture. Science 369, 988–992 (2020).

53. Nieuwenhuys, R., Voogd, J. & van Huijzen, C. The Human Central Nervous System. (Springer Science & Business Media, 2013).

54. Curot, J. et al. Memory scrutinized through electrical brain stimulation: A review of 80 years of experiential phenomena. Neuroscience & Biobehavioral Reviews 78, 161–177 (2017).

55. Ponce, F. A. et al. Bilateral deep brain stimulation of the fornix for Alzheimer’s disease: surgical safety in the ADvance trial. J Neurosurg 125, 75–84 (2016).

56. Aggleton, J. P., Pralus, A., Nelson, A. J. D. & Hornberger, M. Thalamic pathology and memory loss in early Alzheimer’s disease: moving the focus from the medial temporal lobe to Papez circuit. Brain 139, 1877–1890 (2016).

57. Horn, A. et al. Probabilistic conversion of neurosurgical DBS electrode coordinates into MNI space. NeuroImage 150, 395–404 (2017).

58. Poldrack, R. A. Can cognitive processes be inferred from neuroimaging data? Trends in Cognitive Sciences 10, 59–63 (2006).

59. Poldrack, R. A. Inferring Mental States from Neuroimaging Data: From Reverse Inference to Large-Scale Decoding. Neuron 72, 692–697 (2011).

60. Musso, M. et al. A single dual-stream framework for syntactic computations in music and language. NeuroImage 117, 267–283 (2015).

61. Noecker, A. M. et al. StimVisionv2: Examples and Applications in Subthalamic Deep Brain Stimulation for Parkinson’s Disease. Neuromodulation: Technology at the Neural Interface 12, 75–11 (2021).

62. Migliaccio, R. et al. Mapping the Progression of Atrophy in Early- and Late-Onset Alzheimer’s Disease. J Alzheimers Dis 46, 351–364 (2015).

63. Villain, N. et al. Sequential relationships between grey matter and white matter atrophy and brain metabolic abnormalities in early Alzheimer’s disease. Brain 133, 3301–3314 (2010).

64. Horn, A. & Kühn, A. A. Lead-DBS: A toolbox for deep brain stimulation electrode localizations and visualizations. NeuroImage 107, 127–135 (2015).

65. Avants, B., Epstein, C., Grossman, M. & Gee, J. Symmetric diffeomorphic image registration with cross-correlation: Evaluating automated labeling of elderly and neurodegenerative brain. Medical Image Analysis 12, 26–41 (2008).

66. Husch, A., V Petersen, M., Gemmar, P., Goncalves, J. & Hertel, F. PaCER - A fully automated method for electrode trajectory and contact reconstruction in deep brain stimulation. Neuroimage Clin 17, 80–89 (2018).

67. Oxenford, S. et al. Lead-OR: A Multimodal Platform for Deep Brain Stimulation Surgery. 2021.08.09.21261792 https://www.medrxiv.org/content/10.1101/2021.08.09.21261792v1 (2021) doi:10.1101/2021.08.09.21261792.

68. Kim, E. J. et al. Glucose metabolism in early onset versus late onset Alzheimer’s disease: an SPM analysis of 120 patients. Brain 128, 1790–1801 (2005).

69. Hebert, L. E., Weuve, J., Scherr, P. A. & Evans, D. A. Alzheimer disease in the United States (2010-2050) estimated using the 2010 census. Neurology 80, 1778–1783 (2013).

70. Wang, Q. et al. Normative vs. patient-specific brain connectivity in Deep Brain Stimulation. NeuroImage 224, (2021).

71. Li, N. et al. A Unified Functional Network Target for Deep Brain Stimulation in Obsessive-Compulsive Disorder. Biological Psychiatry 90, 701–713 (2021).

72. Jack, C. R. et al. Introduction to the recommendations from the National Institute on Aging-Alzheimer’s Association workgroups on diagnostic guidelines for Alzheimer’s disease. Alzheimers Dement 7, 257–262 (2011).

73. Mohs, R. C. et al. Development of Cognitive Instruments for Use in Clinical Trials of Antidementia Drugs: Additions to the Alzheimer’s Disease Assessment Scale That Broaden Its Scope. Alzheimer Dis Assoc. Disord. 11, S13–S21 (1997).

74. Rosen, W. G., Mohs, R. C. & Davis, K. L. A new rating scale for Alzheimer’s disease. Am J Psychiatry 141, 1356–1364 (1984).

75. Piolino, P., Desgranges, B. & Eustache, F. Episodic autobiographical memories over the course of time: Cognitive, neuropsychological and neuroimaging findings. Neuropsychologia 47, 2314–2329 (2009).

76. Klein, A. et al. Evaluation of 14 nonlinear deformation algorithms applied to human brain MRI registration. NeuroImage 46, 786–802 (2009).

77. Vorwerk, J., Oostenveld, R., Piastra, M. C., Magyari, L. & Wolters, C. H. The FieldTrip-SimBio pipeline for EEG forward solutions. Biomed Eng Online 17, 37 (2018).

78. Pakkenberg, B. & Gundersen, H. J. Neocortical neuron number in humans: effect of sex and age. J Comp Neurol 384, 312–320 (1997).

79. Zalesky, A. & Fornito, A. A DTI-derived measure of cortico-cortical connectivity. IEEE Trans Med Imaging 28, 1023–1036 (2009).

80. Alle, H. & Geiger, J. R. P. Combined analog and action potential coding in hippocampal mossy fibers. Science 311, 1290–1293 (2006).

81. Groppa, S. et al. Physiological and anatomical decomposition of subthalamic neurostimulation effects in essential tremor. Brain 137, 109–121 (2014).

82. Reich, M. M. et al. Short pulse width widens the therapeutic window of subthalamic neurostimulation. Ann Clin Transl Neurol 2, 427–432 (2015).

83. Howell, B. et al. Image-based biophysical modeling predicts cortical potentials evoked with subthalamic deep brain stimulation. Brain Stimulation 14, 549–563 (2021).

84. Wen-Yih Isaac Tseng, F.-C. Y. Generalized q-Sampling Imaging. IEEE TRANSACTIONS ON MEDICAL IMAGING 29, 1626–1635 (2010).

85. Lancaster, J. L. et al. Automated Analysis of Fundamental Features of Brain Structures. Neuroinform 9, 371–380 (2011).

