## Supplementary Material for "Optimal Stimulation Sites and Networks for Deep Brain Stimulation of the Fornix in Alzheimer’s Disease"

*** Corresponding Author**

Andreas Horn, MD, PhD

Associate Professor of Neurology, Center for Brain Circuit Therapeutics, Department of Neurology, Brigham and Women's Hospital, MA, USA.

### Abstract

Deep brain stimulation (DBS) to the fornix is an investigational treatment option for patients with mild Alzheimer's Disease. Outcomes from randomized clinical trials have shown that cognitive function improved in some patients but deteriorated in others. One reason could be variance in electrode placement leading to differential engagement of neural circuits. To investigate this, we analyzed a multi-center cohort of 46 patients with DBS to the fornix. Using normative structural and functional connectivity data, we demonstrate that stimulation of the circuit of Papez and stria terminalis robustly associated with cognitive improvement ($R = 0.45$, $p = 0.031$). On a local level, the optimal stimulation site resided at the direct interface between these structures ($R = 0.33$, $p = 0.016$). Finally, modulating specific distributed brain networks related to memory accounted for optimal outcomes ($R = 0.38$, $p = 0.006$). Findings were robust to multiple cross-validation designs and may now define an optimal network target which could subsequently guide refinement of DBS surgery and programming.

*Table S1. Inclusion/exclusion criteria of Toronto-based pilot clinical trial (NCT00658125)*

| Inclusion Criteria | Exclusion Criteria |
|---|---|
| 1. Man or woman aged 40 to 80 years old | 1. Pre-existing structural brain abnormalities (such as tumor, infarction, or intracranial hematoma) |
| 2. Satisfies the diagnostic criteria for probable AD* | 2. Other neurologic or psychiatric diagnoses |
| 3. Has received the diagnosis of AD within the past 2 years | 3. Medical comorbidities that would preclude patients from undergoing surgery |
| 4. Has a Clinical Dementia Rating (CDR) score of 0.5 or 1.0 | |
| 5. Has a score between 18 and 28 on the Mini Mental State Examination (MMSE) | |
| 6. Has been taking a stable dose of cholinesterase inhibitors for a minimum of 6 months | |

*See McKhann G, Drachman D, Folstein M, et al. Clinical diagnosis of Alzheimer's disease: report of the NINCDS-ADRDA Work Group under the auspices of Department of Health and Human Services Task Force on Alzheimer's Disease. Neurology 1983;34: 939–944.

*Table S2. Inclusion/exclusion criteria of ADvance multi-centre trial (NCT0160806)*

| Inclusion Criteria | Exclusion Criteria |
|---|---|
| 1. 45 years of age (inclusive). | 1. Must meet certain criteria on cognitive and behavioral scales. |
| 2. Probable Alzheimer's disease according to the National Institute of Aging Alzheimer's disease Association criteria. | 2. Current major psychiatric disorder such as schizophrenia, bipolar disorder or major depressive disorder based on psychiatric consult at screening visit. |
| 3. Must meet certain criteria on cognitive and behavioral rating scales. | 3. History of head trauma in the 2 years prior to signing the consent to participate in the study. |
| 4. If female, subjects who are post-menopausal or surgically sterile or willing to use birth control methods for the duration of the study. | 4. History of brain tumor, subdural hematoma, or other clinically significant (in the judgement of the investigator) space-occupying lesion on CT or MRI. |
| 5. An available caregiver willing to participate. | 5. Active psychiatric disorder. |
| 6. Subject is living at home and likely to remain at home for the study duration. | 6. Mental retardation. |

| Inclusion Criteria | Exclusion Criteria |
|---|---|
| 7. The subject is currently taking a stable dose of cholinesterase inhibitor (AChEI) medication for at least 60 days. | 7. Current alcohol or substance abuse as defined by Diagnostic and Statistical Manual of Mental Disorders, Fourth Edition, Text Revision (DSM-IV-TR). |
| | 8. Contraindications for PET scanning (e.g., insulin dependent diabetes). |
| | 9. Contraindications for MRI scanning, including implanted metallic devices (e.g., non-MRI-safe cardiac pacemaker or neurostimulator; some artificial joints metal pins; surgical clips; or other implanted metal parts), or claustrophobia or discomfort in confined spaces. |
| | 10. Abnormal lab results that, in the opinion of the investigator and/or enrollment review committee, would preclude participation in the study. |
| | 11. Abnormal cardiovascular or neurovascular disorder that, in the opinion of the investigator and/or enrollment review committee, would preclude participation in the study. |
| | 12. Unstable doses of any medication prescribed for the treatment of memory loss or Alzheimer's disease. |
| | 13. Currently prescribed any non-AD medications that, in the opinion of the investigator and/or enrollment committee, would preclude participation in the study. |
| | 14. Is unable or unwilling to comply with protocol follow-up requirements. |
| | 15. Has a life expectancy of < 1 year. |
| | 16. Is actively enrolled in another concurrent clinical trial. |

*Table S3. ADAS-cog 11 scores and group assignment of patients. Absolute change calculated subtracting Baseline ADAS-cog 11 from 12-month ADAS-cog 11 value.*

| Patient ID | Baseline ADAS-cog 11 | 12-month ADAS-cog 11 | Absolute change (post-pre) | Relative change (post-pre)/pre | Group |
|---|---|---|---|---|---|
| 01 | 28 | 34 | 6 | 21.42 | Poor responders |
| 02 | 22 | 30 | 8 | 36.36 | Poor responders |
| 03 | 19 | 34 | 5 | 78.94 | Poor responders |
| 04 | 17 | 39 | 22 | 129.41 | Poor responders |
| 05 | 19 | 21 | 2 | 10.52 | Middle responders |
| 06 | 13 | 18 | 5 | 38.46 | Poor responders |
| 07 | 13 | 15 | 2 | 15.38 | Middle responders |
| 08 | 24 | 31 | 7 | 29.16 | Poor responders |
| 09 | 23 | 30 | 7 | 30.43 | Poor responders |
| 10 | 13 | 24 | 11 | 84.61 | Poor responders |
| 11 | 12 | 7 | -5 | -41.67 | Top responders |
| 12 | 15 | 24 | 9 | 60 | Poor responders |
| 13 | 31 | 36 | 5 | 16.13 | Middle responders |
| 14 | 29 | 43 | 14 | 48.28 | Poor responders |
| 15 | 19 | 26 | 7 | 36.84 | Poor responders |
| 16 | 32 | 33 | 1 | 3.13 | Middle responders |
| 17 | 16 | 29 | 13 | 81.25 | Poor responders |
| 18 | 18 | 23 | 5 | 27.78 | Poor responders |
| 19 | 23 | 24 | 1 | 4.35 | Middle responders |
| 20 | 15 | 36 | 11 | 140 | Poor responders |
| 21 | 22 | 10 | -12 | -54.55 | Top responders |
| 22 | 16 | 19 | 3 | 18.75 | Middle responders |
| 23 | 16 | 22 | 6 | 37.5 | Poor responders |
| 24 | 21 | 42 | 21 | 100 | Poor responders |
| 25 | 17 | 30 | 13 | 76.48 | Poor responders |
| 26 | 24 | 29 | 5 | 20.83 | Middle responders |
| 27 | 28 | 38 | 10 | 35.71 | Poor responders |

| Patient ID | Baseline ADAS-cog 11 | 12-month ADAS-cog 11 | Absolute change (post-pre) | Relative change (post-pre)/pre | Group |
|---|---|---|---|---|---|
| 28 | 14 | 15 | 1 | 7.14 | Middle responders |
| 29 | 20 | 28 | 8 | 40 | Poor responders |
| 30 | 16 | 15 | -1 | -6.25 | Top responders |
| 31 | 17 | 12 | -5 | -29.41 | Top responders |
| 32 | 35 | 51 | 16 | 45.71 | Poor responders |
| 33 | 22 | 39 | 17 | 72.27 | Poor responders |
| 34 | 21 | 23 | 2 | 9.52 | Middle responders |
| 35 | 17 | 35 | 18 | 105.88 | Poor responders |
| 36 | 22 | 18 | -4 | -18.18 | Top responders |
| 37 | 19 | 19 | 0 | 0 | Middle responders |
| 38 | 18 | 17 | -1 | -5.56 | Top responders |
| 39 | 16 | 15 | -1 | -6.25 | Top responders |
| 40 | 13 | 21 | 8 | 61.54 | Poor responders |
| 41 | 18 | 17 | -1 | -5.56 | Top responders |
| 42 | 11 | 17 | 6 | 54.55 | Poor responders |
| 43 | 21 | 22 | 1 | 4.76 | Middle responders |
| 44 | 13 | 40 | 27 | 207.69 | Poor responders |
| 45 | 10 | 19 | 9 | 90 | Poor responders |
| 46 | 22 | 19 | -3 | -13.64 | Top responders |

### Normative Connectomes: Underlying Data

*Table S4. Specification of normative connectome data. Abbreviations: TR = Repetition time, TE = Echo time, FOV = Field of view, BOLD = Blood oxygenation level-dependent, EPI = Gradient-echo echo-planar imaging, FA = Flip angle*

| Connectome | Scan parameters | References | Data sources |
|---|---|---|---|
| Structural: *In vivo* human whole-brain Connectom diffusion MRI dataset at 760 μm istotropic resolution | Scanner: MGH-USC 3T Connectom.<br>Maximum gradient strength of 300mT/m and maximum slew rate of 200 T/m/s, custom-built 64-channel phased-array coil.<br>gSlider-SMS sequence.<br>gSlider encoding: 5<br>MB factor: 2<br>$R_{inplane}$ factor: 3<br>Acquisition: Axial (PE along AP/PA)<br>TR/TE: 3500/75 ms<br>FOV: 220.0 ∗ 218.5 mm<br>Acquisition matrix: 290 ∗ 288<br>Acquired slices: 190<br>Slice thickness: 0.76 mm<br>Effective echo spacing: 0.34 ms<br>Readout bandwidth: 1150 Hz/Pixel<br>Phase partial Fourier: 6/8<br>b-values: 1000, 2500 s/mm$^2$<br>144 (b0), 420 (b1000), 840 (b2500) w/AP/PA (total 2808 volumes)<br>Total acquisition time: ~14.5 hours | Wang et al. (2021) Sci. Data[1] | 9 two-hour scan sessions<br>1 healthy subject |
| **Functional**: The organization of the human cerebral cortex estimated by intrinsic functional connectivity | Scanner: 3T Tim Trio scanners (Siemens, Erlangen, Germany)<br>12-channel receive coil array, Gradient -echo echo-planar imaging (EPI) sequence sensitive to BOLD contrast.<br>Acquisition: Slices aligned to anterior commissure-posterior commissure plane<br>EPI parameters<br>TR/TE: 3000 ms/30 ms<br>FA: 85°, 3 * 3 * 3-mm voxels<br>FOV: 216<br>47 axial slices collected with intervaled acquisition, no gap between slices<br>6.2 minute-functional run (124 timepoints) | Yeo et al. (2011) J. Neurophysiol[2]<br><br>Holmes et al. (2015) Sci. Data[3] | Resting-state fMRI data from 1,000 health subjects (average 1.7 runs per subject) |

**Final model parameters of DBS fiber filtering**

During the training phase of model optimization in the DBS fiber filtering analysis, a variety of parameters were tested with the aim to create a tract-set that was i) robustly predictive during cross-validation within the training set (leave-one-out and several k-fold strategies were interactively tested) and ii) was *not robust* to permutations of improvement data (also see fig. 4). The following set of parameters were finally selected and used to cross-predict outcomes in the test-cohort.

*Table S5. Model parameters available in the DBS fiber filtering tool implemented in Lead-DBS, their units, range, and selected value, as well as a brief explanation of what the parameter means.*

| Model parameter | Unit | Range | Selected Value | Explanation |
|---|---|---|---|---|
| Model Setup | N/A | [Sum, Mean, Peak, 5% Peak] | Peak | The peak value of each E-field and each Tract was considered |
| Correlation Type | N/A | [Pearson, Spearman, Bend] | Spearman | Spearman's rank correlations were used (see methods: Model considerations) |
| Tracts "connected" if peak E-field magnitude they traverse is above | V/m | 0.05 – 2.5 | 0.36 V/m | (see next parameter) |
| Tracts must be connected to > x % of E-fields | % | 0 – 100 | 20% | Tracts were only considered if they traversed regions with > 0.36 V/m in > 20 % E-fields |
| Show/Use number of Fibers | % | 0 – 100 | 70% | 70% of fibers with positive R-values were visualized and used in predictive models (cross-validations within training cohort and training > test cross-predictions) |
| Base Prediction On | N/A | [Sum, Mean, Peak, 5% Peak, Profile of Scores: Pearson, Profile of Scores: Spearman, Profile of Scores: Bend] | Profile of Scores: Spearman | This setting was used since it follows the same logic as in sweetspot and network mapping approaches also used here. |

### Sweetspot Coordinates

*Table S6. Probabilistic Stimulation Mapping peak and center coordinates in non-mirrored and mirrored data.*

| | **Sweetspot** | | **Sourspot** | |
|---|---|---|---|---|
| | *Unflipped analysis* | | | |
| | **LH: 552 voxels, 14.9 mm³** | **RH: 267 voxels, 7.09 mm³** | **LH: 516 voxels, 13.93 mm³** | **RH: 892 voxels, 24.08 mm³** |
| **Peak coordinate** | X = -3.9 mm<br>Y = -1.5 mm<br>Z = -3.6 mm | X = 3 mm<br>Y = -0.9 mm<br>Z = -3 mm | X = -5.4 mm<br>Y = -0.3 mm<br>Z = -6.9 mm | X = 4.5 mm<br>Y = 2.1 mm<br>Z = -4.2 mm |
| **Cluster center** | X = -5.1 mm<br>Y = 0.9 mm<br>Z = -3.3 mm | X = 2.4 mm<br>Y = -0.3 mm<br>Z = -3 mm | X = -4.2 mm<br>Y = 0 mm<br>Z = -6.9 mm | X = 3.9 mm<br>Y = 0.9 mm<br>Z = -5.7 mm |
| | *Flipped Analysis* | | | |
| | **LH: 392 voxels, 10.58 mm³** | **RH: 586 voxels, 15.82 mm³** | **LH: 734 voxels, 19.81 mm³** | **RH: 797 voxels, 21.51 mm³** |
| **Peak coordinate (fig. 4)** | X = -3.9 mm<br>Y = -1.5 mm<br>Z = -3 mm | X = 6.9 mm<br>Y = 0 mm<br>Z = -5.1 mm | X = -5.1 mm<br>Y = 2.4 mm<br>Z = -4.8 mm | X = 1.5 mm<br>Y = 0.3 mm<br>Z = -6.9 mm |
| **Cluster center (fig. 4)** | X = -4.8 mm<br>Y = -0.9 mm<br>Z = -3.6 mm | X = 3.9 mm<br>Y = 1.2 mm<br>Z = -3.3 mm | X = -4.5 mm<br>Y = 0 mm<br>Z = -6.6 mm | X = 3.6 mm<br>Y = 0.3 mm<br>Z = -6.3 mm |

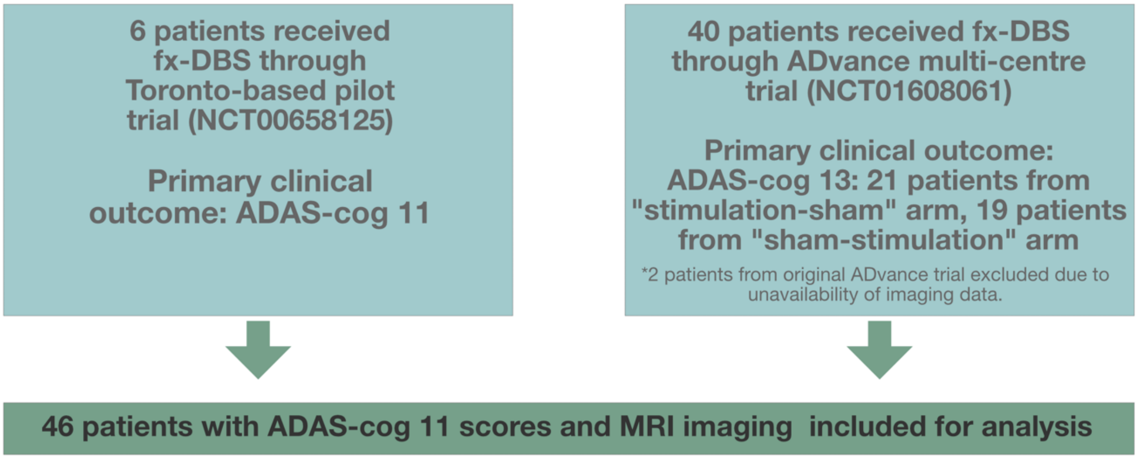


*Figure S1. Flowchart explaining patient inclusion for this work.*

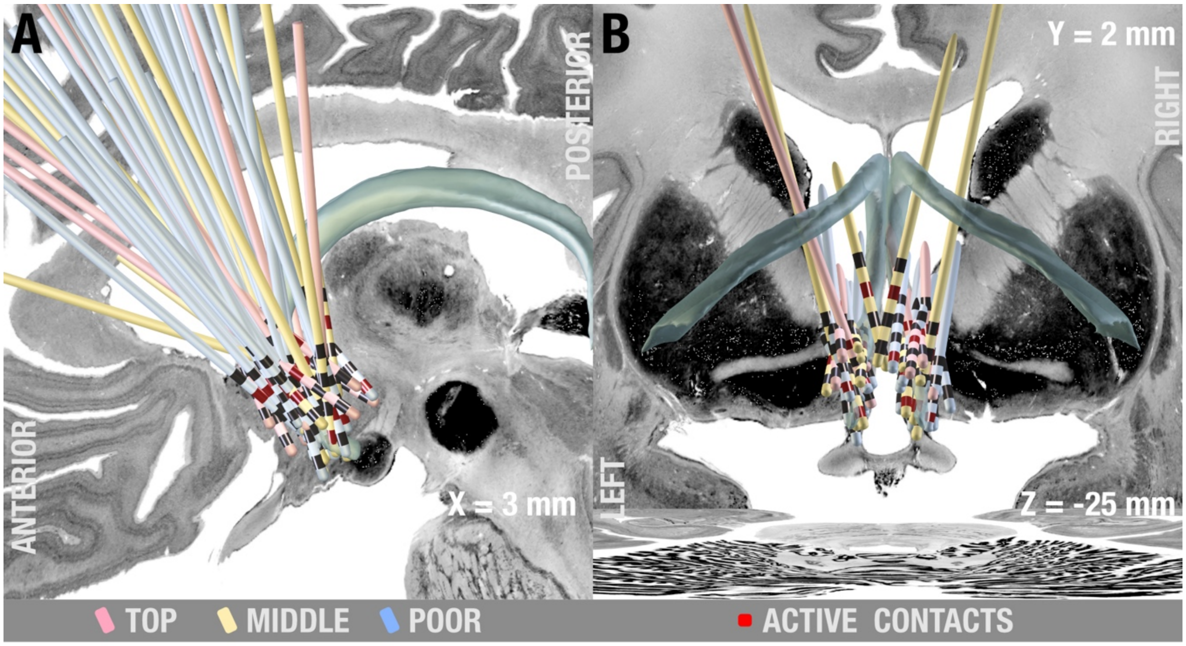


***Figure S2.*** *Electrode localizations. A) Sagittal and B) coronal view showing solid electrodes classified by outcome group (blue-poor responders, yellow-middle responders, pink-top-responders), active contacts highlighted with red superimposed on slices of a brain cytoarchitecture atlas in MNI 152 space*[4]*. Fornix informed by the CoBrALab Atlas* [5]*.*

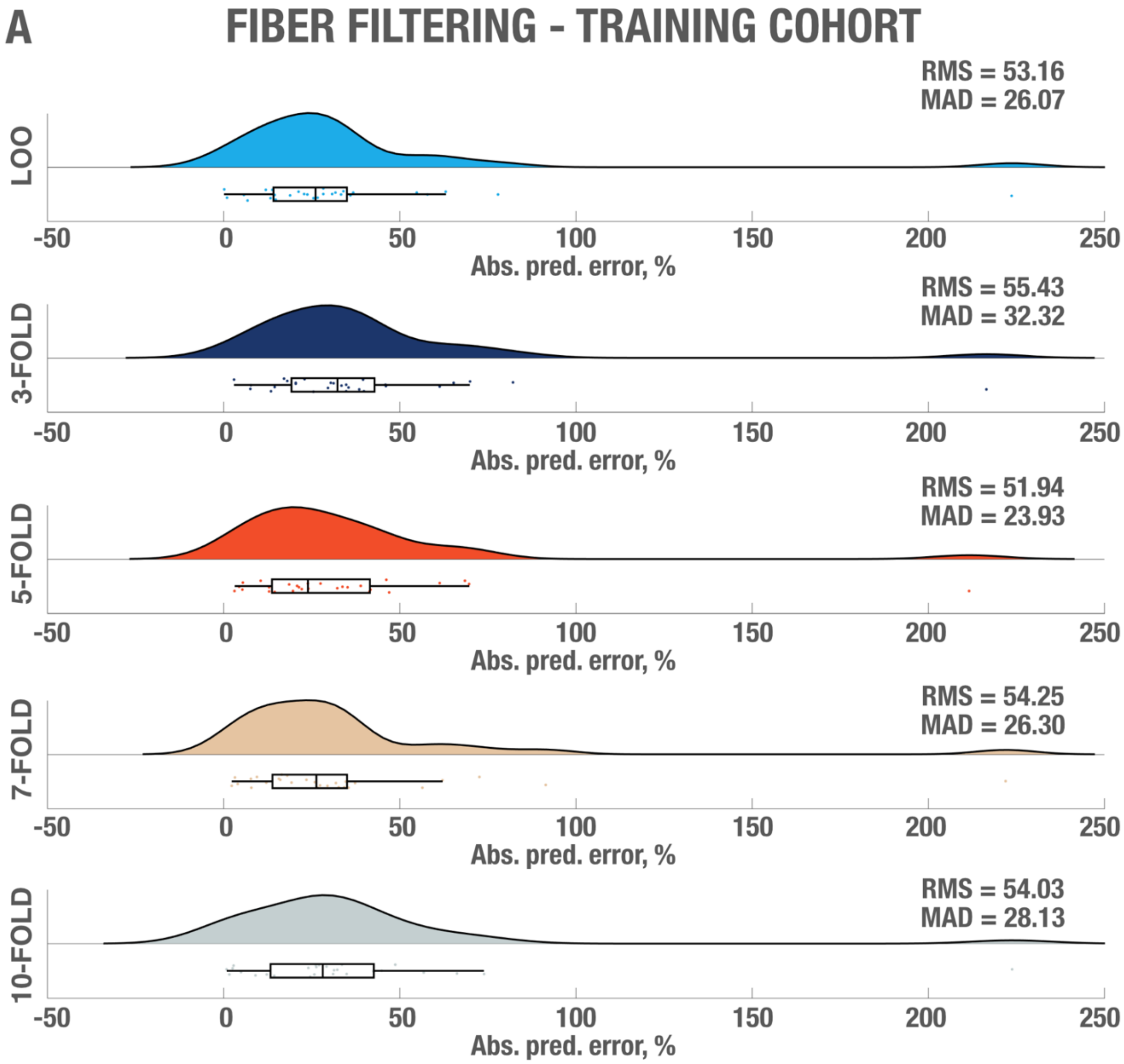


*Figure S3. In-fold analysis from fiber filtering analysis on Training cohort showing absolute predicted error, root mean square deviation (RMS) and median absolute deviation (MAD) for each of the validation approaches.*

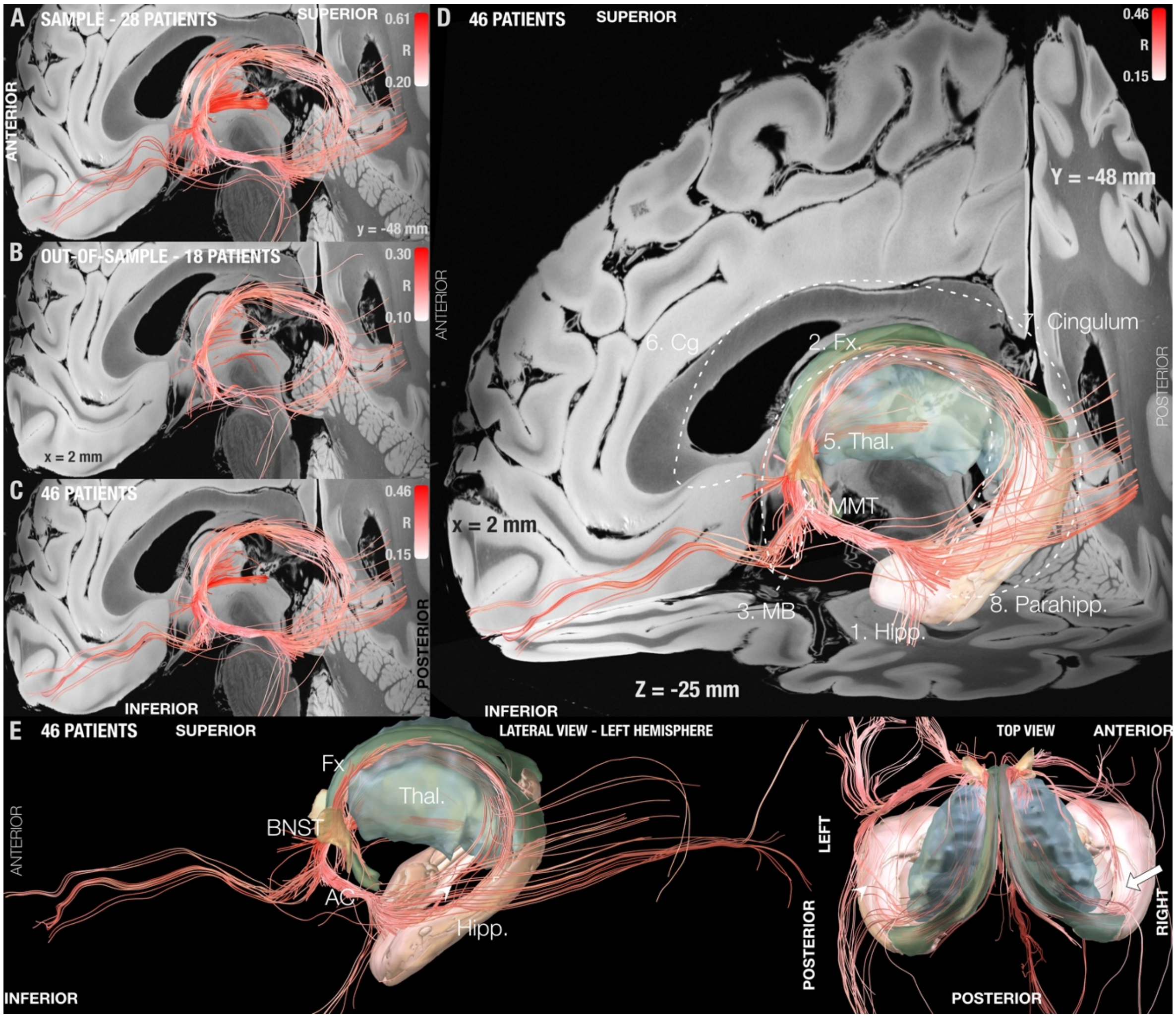


***Figure S4.*** *Fiber tracts associated with optimal clinical response superimposed on slices of a 100-µm, 7T brain scan in MNI 152 space. From a set of 5 million fiber tracts sampled from a high-resolution connectome, the ones preferably modulated by top-responding (and not by poor-responding) patients were selected using the DBS fiber filtering method and visualized. The process was repeated on the training-cohort (N = 30) (A), the test-cohort (N = 20) (B), and both cohorts combined (N = 50) (C). Fiber tracts are color-coded by the resulting Spearman's rank correlation coefficients which shows how strongly modulating each bundle correlated with clinical response across patients. D) Results from panel C superimposed on atlas structures forming part of the circuit of Papez, also visualized by dotted arrows. E) Lateral and top views of fibertract superimposed with structures of interest, white arrow indicates intersection of streamlines of the fornix and AC that could give the illusion of a loop on lateral projection views. 1. Hipp = Hippocampus, 2. Fx. = Fornix, 3. MB = mamillary bodies, 4. MMT = mamillothalamic tract, 5. Thal. = thalamus, 6. Cg Cingulate gyrus, 7. Cingulum and 8. Parahipp: Parahippocampal gyrus. The backdrop features an ultra-high resolution (100 µm) template of the human brain[6]. Structures: Fornix (blue-green), Hippocampus (pink), Thalamus(blue) informed by the CoBrALab Atlas[5], Bed nucleus of the stria terminalis (light brown) informed by the Atlas of the Human Hypothalamus[7].*

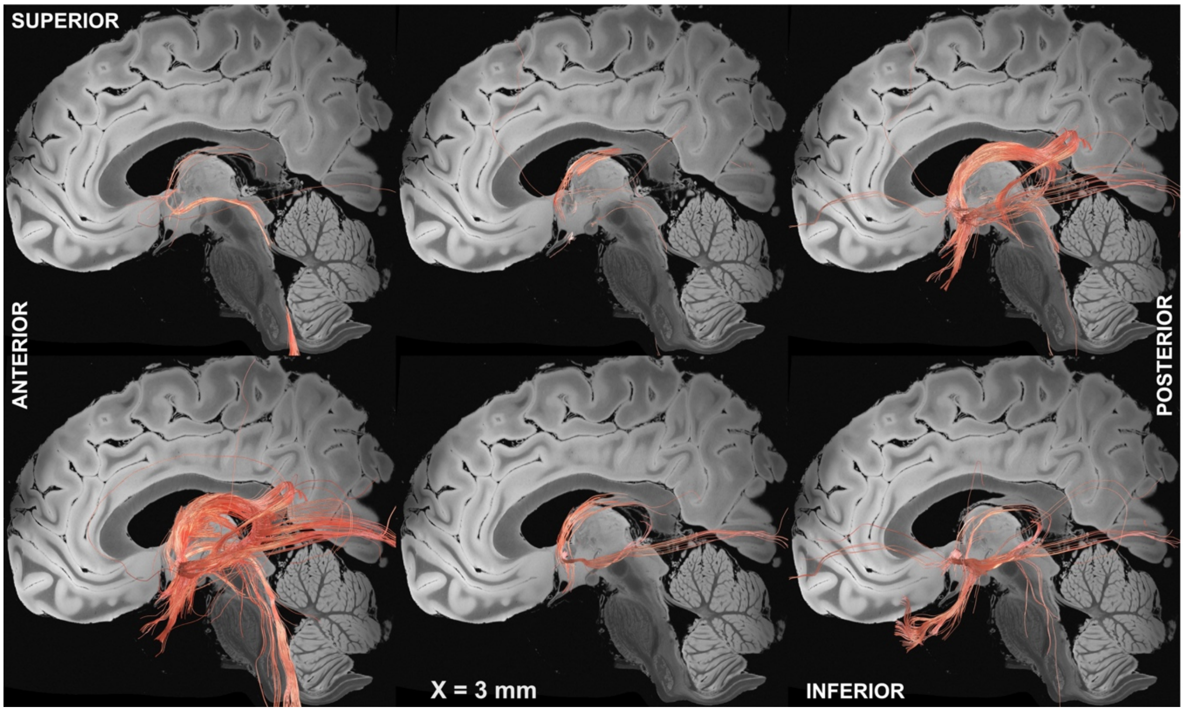


***Figure S5.*** *Random permutation results. Example results when repeating the DBS fiber filtering method after randomly permuting clinical improvement values across patients. This analysis was performed to demonstrate that tract results do not merely reflect average connectivity of the group of DBS electrodes but are highly informed by improvement values. For instance, the result on the top left highlights a connection to the brainstem, the one in the top middle the stria terminalis and the one on the bottom middle and right panels the anterior commissure.*

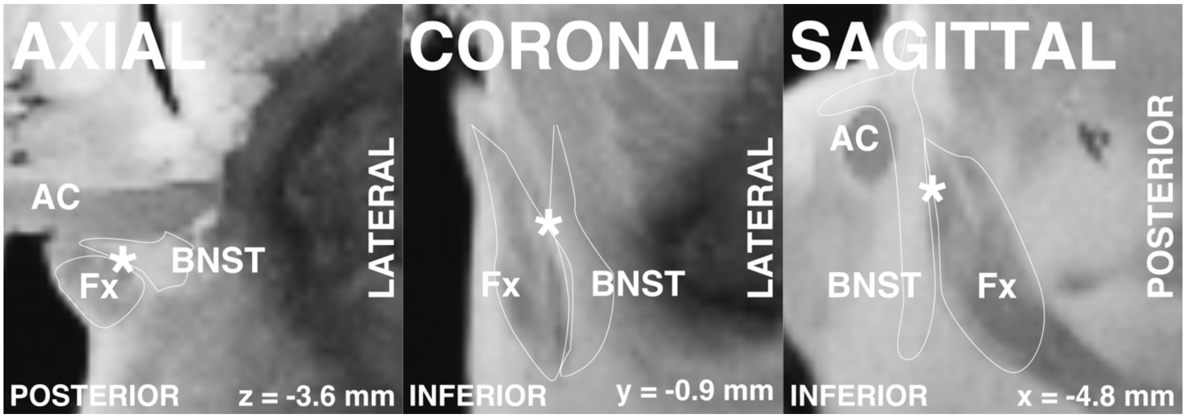


***Figure S6.*** *Close up view of positive sweetspot cluster center coordinate at the junction between fornix and bed nucleus of stria terminalis (BNST). Fx: Fornix, AC: Anterior Commissure.*

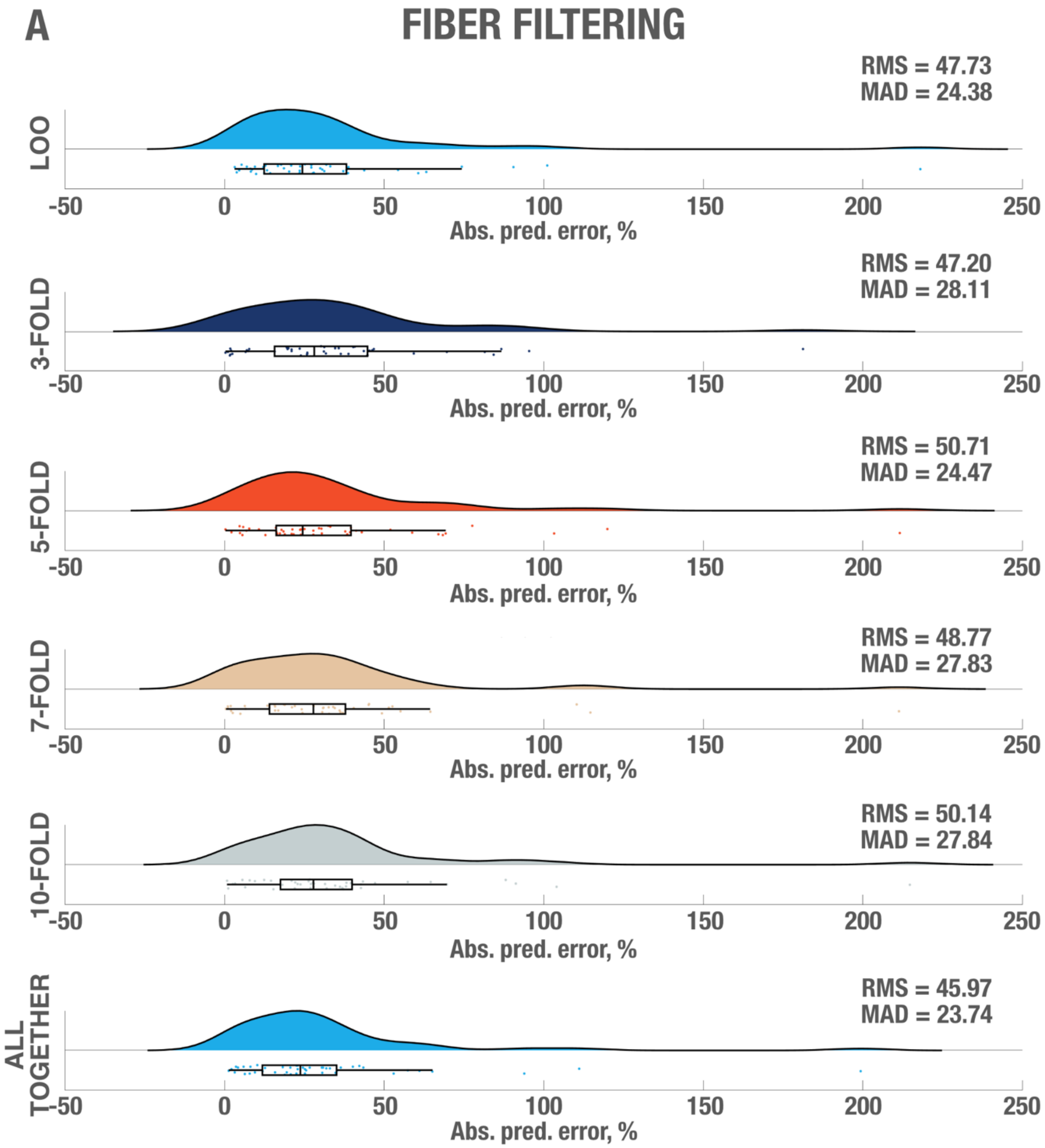
A
FIBER FILTERING
RMS = 47.73
MAD = 24.38
LOO
-50 0 50 100 150 200 250
Abs. pred. error, %
RMS = 47.20
MAD = 28.11
3-FOLD
-50 0 50 100 150 200 250
Abs. pred. error, %
RMS = 50.71
MAD = 24.47
5-FOLD
-50 0 50 100 150 200 250
Abs. pred. error, %
RMS = 48.77
MAD = 27.83
7-FOLD
-50 0 50 100 150 200 250
Abs. pred. error, %
RMS = 50.14
MAD = 27.84
10-FOLD
-50 0 50 100 150 200 250
Abs. pred. error, %
RMS = 45.97
MAD = 23.74
ALL TOGETHER
-50 0 50 100 150 200 250
Abs. pred. error, %

*Continuation Figure S7*

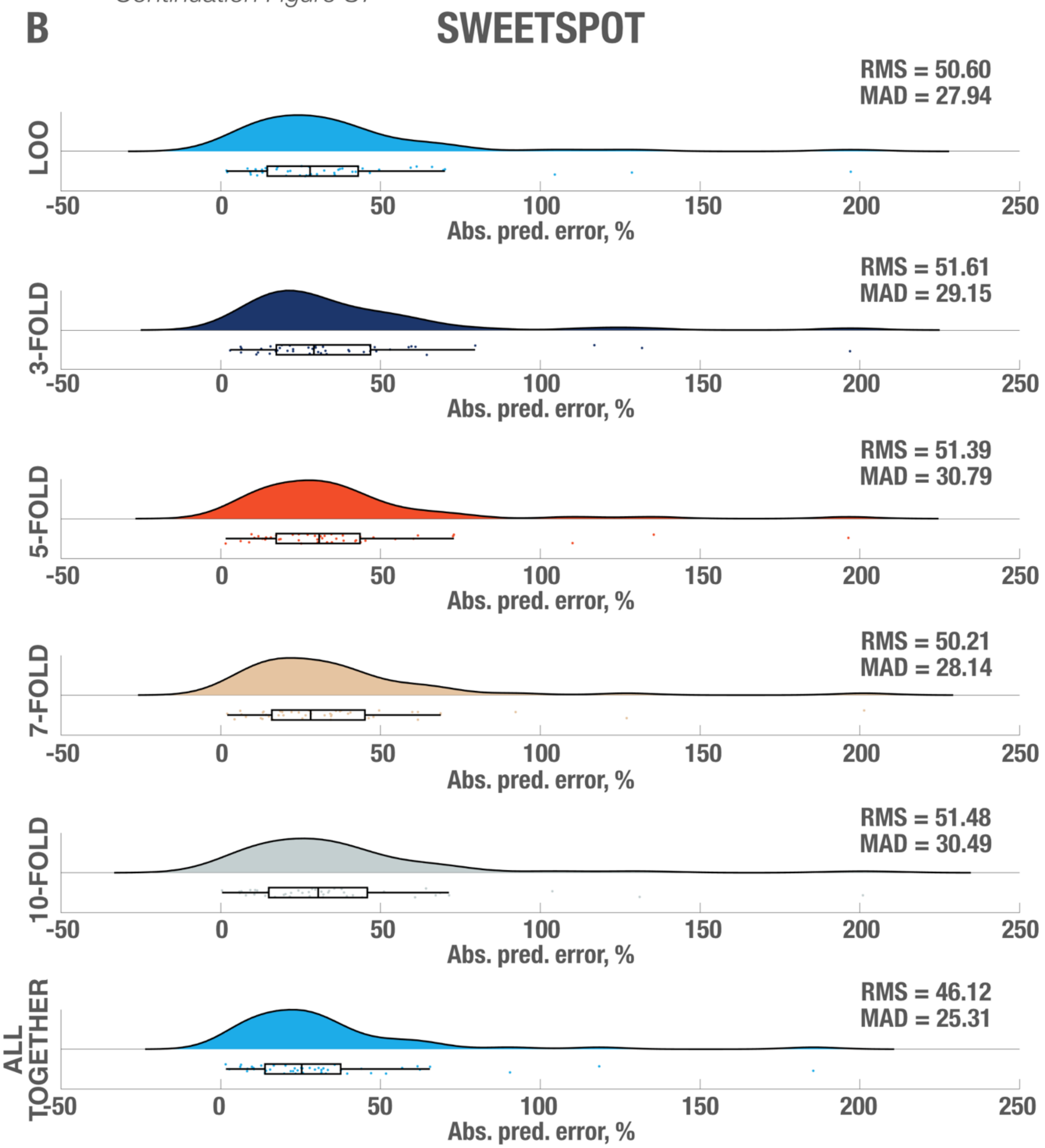

*Continuation Figure S7*

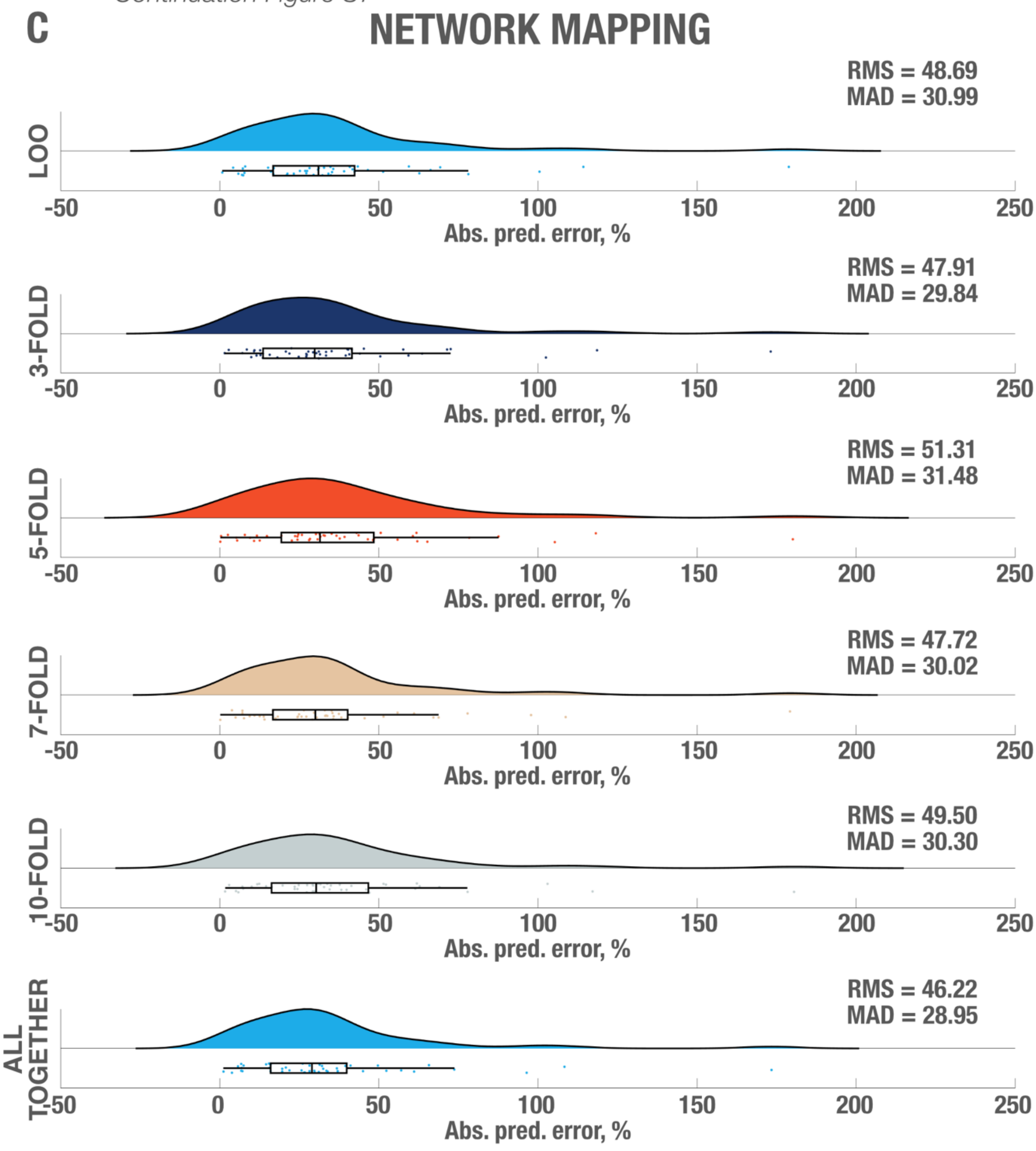


*Figure S7. In-fold analysis from summary showing absolute predicted error, root mean square deviation (RMS) and median absolute deviation (MAD) for each of the validation approaches followed on fiber filtering (A), sweetspot (B) and network mapping (C) methods.*

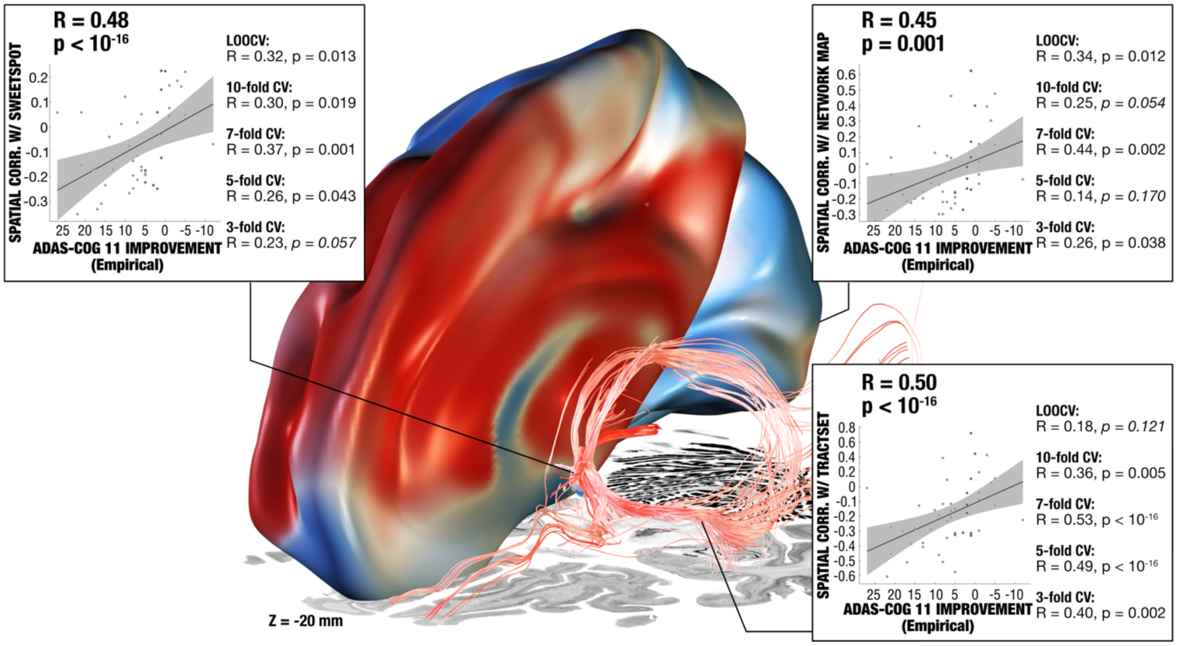


***Figure S8.*** *Results summary including the models from sweetspot, tract- and network-levels calculated with absolute ADAS-cog 11 outcomes. The three levels of analysis led to mostly significant predictions of clinical outcomes across leave-one-patient-out and multiple k-fold designs. Gray shaded areas represent 95% confidence intervals.*

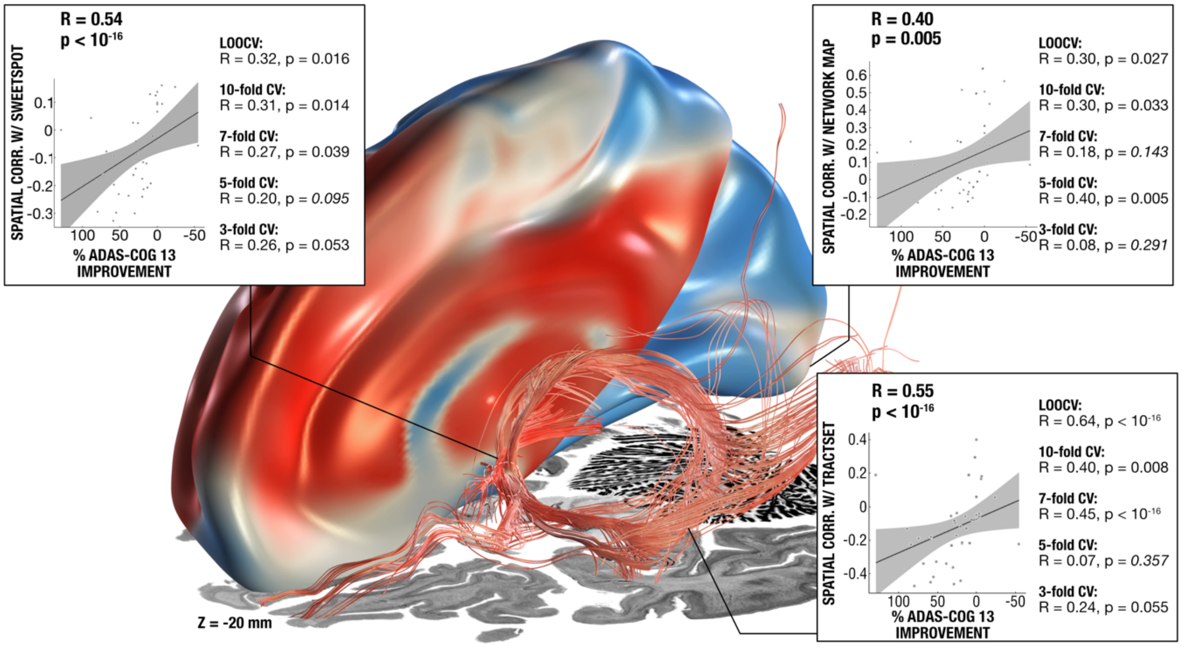


***Figure S9.*** *Results summary including the models from sweetspot, tract- and network-levels calculated with ADAS-cog 13 outcomes (Note: only participants of ADvance trial were included for this subanalysis, N = 40). The three levels of analysis led to mostly significant predictions of clinical outcomes across leave-one-patient-out and multiple k-fold designs. Gray shaded areas represent 95% confidence intervals.*

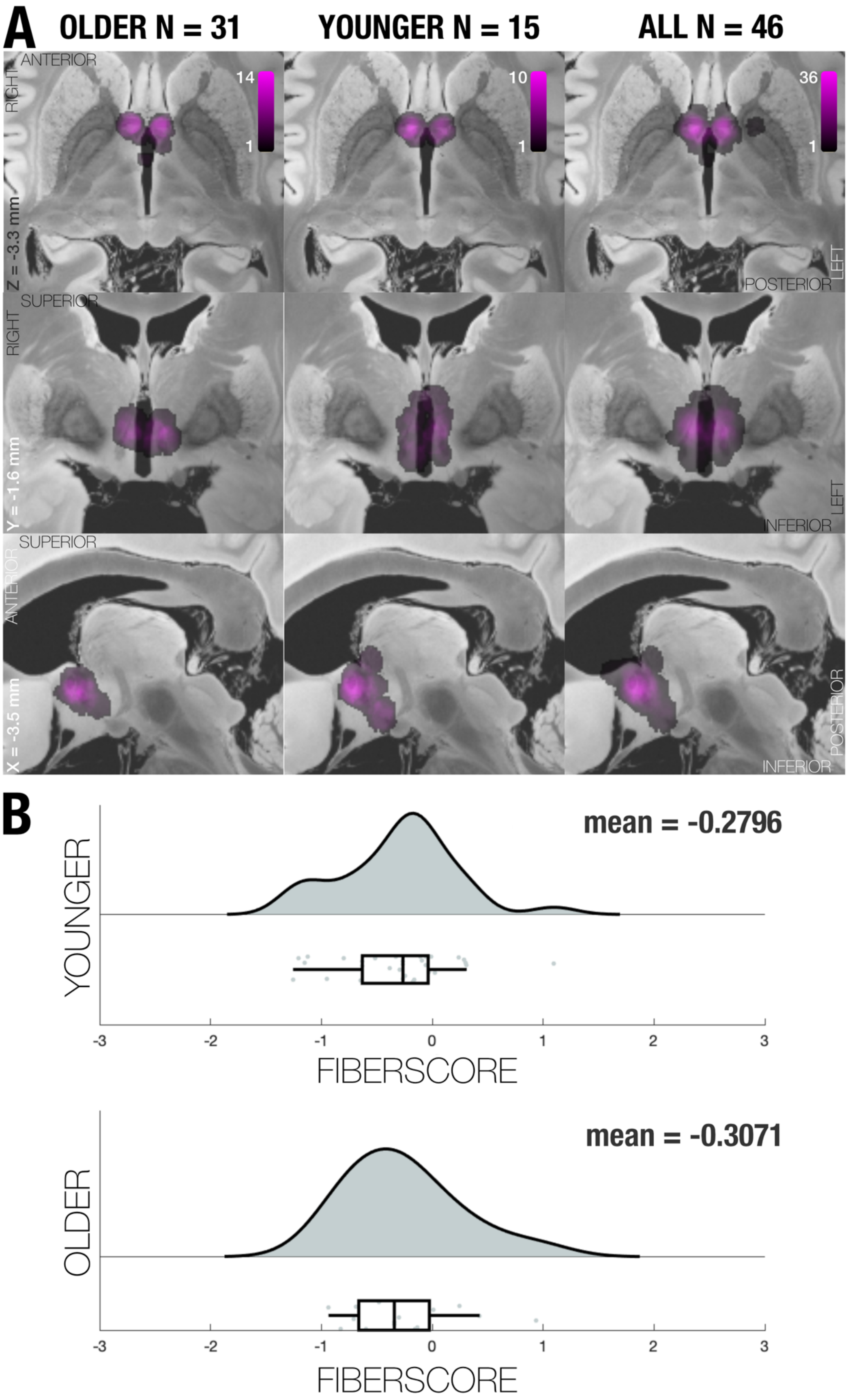

A
OLDER N = 31
YOUNGER N = 15
ALL N = 46
ANTERIOR
RIGHT
Z = -3.3 mm
14
1
10
1
36
1
POSTERIOR
LEFT
SUPERIOR
RIGHT
Y = -1.6 mm
INFERIOR
LEFT
SUPERIOR
ANTERIOR
X = -3.5 mm
POSTERIOR
INFERIOR
B
YOUNGER
mean = -0.2796
-3
-2
-1
0
1
2
3
FIBERSCORE
OLDER
mean = -0.3071
-3
-2
-1
0
1
2
3
FIBERSCORE

***Figure S10**. Effects of Age. A) Axial, coronal, and sagittal overlay of maps created from stimulation volumes of subjects older than 65 years (left), younger than 65 years (middle) and whole cohort (right). B) Fiberscores obtained through DBS fiber filtering analysis explained in Methods and Results sections, by the stimulation volumes of younger than 65-year-old patients (top), and patients 65-year-old or older (bottom), p(T-test) = 0.790. The model used to estimate these scores was calculated in a leave-one-patient out design across the entire cohort.*

**Supplementary Methods**

Narrative section of methods / predictive models:

In all three models, each patient contributed their relative improvement of ADAS-cog-11 scores (before surgery, one year after surgery).

Beyond that, each model (i) tracts, ii) sweetspots and iii) functional networks) was run independently from one another.

- i) For tracts, each patient contributed the peak E-field amplitude that each tract of the normative connectome was modulated by.
- ii) For sweetspots, each patient contributed the modeled electric field in MNI space (represented as a NIfTI volume).
- iii) For functional networks, each patient contributed a (normative) rs-fMRI map seeding from the individual patient ("connectivity fingerprints").

Then, the three models created a i) combination of tracts ii) optimal target (sweetspot), and iii) functional network profile associated with optimal clinical improvements.

- i) For tracts, this was achieved by rank correlating the modulation amplitude imposed on each tract with clinical improvements across the set of patients. This led to an R-value for each tract, denoting how well its modulation correlated with clinical improvements (the concept was introduced in Irmen et al. 2019 Annals of Neurology).
- ii) For sweetspots, this was achieved by rank correlating each voxel with clinical outcomes across the set of patients. This led to an R-map denoting how well modulations of specific voxels correlated with clinical outcomes (the concept was introduced in Horn et al. 2022 PNAS).
- iii) For functional networks, this was achieved by correlating the voxel values of connectivity fingerprints with clinical improvements across the set of patients. This led to an R-map denoting how well connectivity estimates between stimulation sites and each voxel in the brain correlated with clinical outcomes (the concept was introduced in Horn et al. 2017 Annals of Neurology).
-

Finally, data was cross-validated within the three models:

- i) For tracts, this was achieved by rank correlating the impacts of the E-Fields of an unseen patient on all tracts and their R-values This led to a fiberscore denoting how specifically an unseen E-Field modulated tracts associated with optimal outcomes (the concept was introduced in Horn et al. 2022 PNAS).

- ii) For sweetspots, this was achieved by spatially correlating the E-Fields of an unseen patient with the R-map model. This led to a sweetspot score denoting correlation coefficients of agreement between the actual stimulation field and an "optimal" stimulation field (represented by the R-map; the concept was introduced in Horn et al. 2022 PNAS).
- iii) For functional networks, this was achieved by spatially correlating the functional connectivity fingerprints with the R-map model. This led to a network score denoting correlation coefficients of agreement between the actual network profile and an optimal network profile (represented by the R-map; the concept was introduced in Horn et al. 2017 Annals of Neurology).
